# Inhospital Mortality, Readmission, and Prolonged Length of Stay Risk Prediction Leveraging Historical Electronic Health Records

**DOI:** 10.1101/2024.04.15.24305875

**Authors:** Rajeev Bopche, Lise Tuset Gustad, Jan Egil Afset, Birgitta Ehrnström, Jan Kristian Damås, Øystein Nytrø

**Affiliations:** Department of Computer Science, Norwegian University of Science and Technology, Trondheim, Norway; Faculty of Nursing and Health Sciences, Nord University, Levanger, Norway; Department of Medical Microbiology, St. Olavs Hospital, Trondheim University Hospital, Trondheim, Norway; Department of Clinical and Molecular Medicine, Norwegian University of Science and Technology, Trondheim, Norway; Department of Computer Science, The Arctic University of Norway, Tromsø. Norway; Department of Infectious Diseases, Clinic of Medicine, St Olavs Hospital, Trondheim, Norway; Clinic of Anaesthesia and Intensive Care, St Olavs Hospital, Trondheim University Hospital, Trondheim; Department of Medicine and Rehabilitation, Levanger Hospital, Nord-Trøndelag Hospital Trust

**Keywords:** Healthcare Informatics, Electronic Patient Records, Tree-based Models, Predictive Analytics, Machine Learning, eXplainable Artificial Intelligence, Mortality, Readmission, Prolonged Length of Stay, Medical History

## Abstract

**Objective:** The aim of this study was to investigate predictive capabilities of historical records of patients maintained at hospitals towards predicting an impending adverse outcomes such as, mortality, readmission, and prolonged length of stay (PLOS).

**Methods:** Leveraging a de-identified dataset from a tertiary care university hospital, we developed a eXplainable Artificial Intelligence (XAI) framework combining tree-based and traditional ML models with interpretations, and statistical analysis of predictors of mortality, readmission, and PLOS.

**Results:** Our framework demonstrated exceptional predictive performance with notable Area Under the Receiver Operating Characteristic (AUROC) of 0.9625 and Area Under the Precision-Recall Curve (AUPRC) of 0.8575 for 30-day mortality at discharge and an AUROC of 0.9545 and AUPRC of 0.8419 at admission. For the readmission and PLOS risk the highest AUROC achieved were 0.8198 and 0.9797 repectively. The tree-based machine learning (ML) models consistently outperformed the traditional ML models in all the four prediction tasks. The key predictors were age, derived temporal features, routine laboratory tests, and diagnostic and procedural codes.

**Conclusion:** The study underscores the potential of leveraging medical history for enhanced predictive analytics in hospitals. We present a accurate and intuitive framework for early warning models that can be easily implemented in the current and developing digital health platforms to accurately predict adverse outcomes.

## 1. INTRODUCTION

Important healthcare indicators, such as 30-day mortality, 30-day readmissions, and Prolonged Length of Stay (PLOS) are essential for managing patient care and allocating resources efficiently,[1–3]. Accurate forecasts of these indicators are pivotal for early identification of high-risk patients, leading to timely medical actions and improved patient outcomes,[4, 5]. An Electronic Health Record (EHR) is a digital repository encompassing a patient’s complete health history, documented from multiple healthcare encounters across various healthcare providers over time,[6]. EHR has been used for secondary applications like disease progression modeling,[7], patient trajectory modeling,[8], disease inference,[9], risk stratification, and survival prediction,[10]. These data-driven analyses are increasingly needed in all kinds of health services and research. However, the problem with EHRs is mostly the sparseness and context-dependent interpretation of EHRs can cause incompleteness and to a lesser extent inconsistency and inaccuracy,[11].

The recent introduction of eXplainable AI (XAI) in health informatics offers transformative potential,[12]. It allows for complex data within EHRs to be analyzed in a way that can be understood by healthcare professionals, supporting informed clinical decisions,[13, 14]. Despite the potential benefits, the application of comprehensive data available within EHRs for predictive analysis is not yet commonplace in hospital settings,[15]. Preliminary research suggests significant healthcare interactions can precede critical health events, such as cancer diagnoses,[16]. There has been no studies that has leveraged the complete information from the medical history of patients and most only consider predictors from a specified time window,[17, 18]. We theorize that historical EHRs contains patterns and early indicators of impending adverse hospital outcomes. To explore this, we conduct retropective analysis of patient medical histories, introducing an XAI-based Risk Analysis and Interpretation (XRAI) framework for prediction of the risk of hospital adverse events: mortality, readmission, and PLOS.

## 2. RELATED WORK

The domain of health informatics (HI) has experienced significant advancements with the integration of artificial intelligence (AI) techniques, especially in predictive analytics. Various AI algorithms have been applied in HI, ranging from traditional machine learning models to more complex deep learning architectures,[19, 20]. Tree-based models have gained popularity due to their robustness in handling medical data,[21]. While AI models can produce accurate predictions, their “black box” nature has been a concern in the medical domain due to the critical nature of healthcare decisions,[22]. This has led to the rise of XAI techniques for local and global model explanations. Tools like SHapley Additive exPlanations (SHAP) and Local Interpretable Model-agnostic Explanations (LIME) have been utilized to provide insights into model decision-making processes,[23, 24]. The significance of XAI in healthcare is profound as it ensures both healthcare practitioners and patients can trust and understand AI-driven decisions,[25]. Though predictive analytics have shown promise, they also come with challenges,[26]. Data quality, missing values, and class imbalances have been cited as some of the significant challenges in healthcare predictions,[27, 28]. Additionally, the ethical implications concerning patient data security and consent are of paramount importance,[29].

Previous work on leveraging longitudinal medical data by Chicco et al. (2020) showed that traditional ML models predicted survival of the patients diagnosed with sepsis using minimal clinical records of patients,[10]. Other works focused on visualizing the medical history or building patient disease trajectories,[30]. Some studies have worked on tackling the problems with the representation of medical data and codes. For example, Tran et. al. (2015) worked on building a low-dimensional representation of medical events using a modified Restricted Boltzmann Machine (RBM). Thereafter they trained a logistic regression classifier for suicide risk stratification,[31]. Similarly, Choi et. al. (2015) disease-specific applications have treated the medical history as a sequence of events and then trained ML models to predict diagnostic outcome in the next event,[32]. While Jia et al. (2020) used patient similarity-based frameworks to group similar patient histories together,[33]. In works predicting adverse outcomes in hospitals, study by Cai et al. (2016),[17] developed a non-disease-specific Bayesian Network model to predict mortality, readmission, and length of stay (LoS) from EHRs. Utilizing data from 32,634 patients admitted via emergency department to a Sydney hospital between 2008 and 2011, the model achieved an average daily accuracy of 80% with an area under the receiver operating characteristic curve (AUROC) of 0.82 for mortality, which was the most predictable outcome. Tavakolian et al. (2023) introduced an optimized hybrid deep model termed Genetic Algorithm-Optimized Convolutional Neural Network (GAOCNN) for predicting hospital readmission and LoS. Tested on three distinct healthcare datasets, this model achieved impressive prediction accuracies, reaching 97.2% for hospital readmission prediction in diabetic patients, and 89.0%, 99.4%, and 94.1% for LoS prediction in diabetic, COVID-19, and ICU patients respectively,[18]. Clark et al. (2016) explored a multistate model for predicting mortality, LoS, and readmission for surgical patients using the American College of Surgeons National Surgical Quality Improvement Program data,[34]. Most model focused on recent data pertaining to the patients and none of the study utilized the predictors from the complete medical history of their patients, apart from demographic and information on co-morbidities. In our previous work through feature engineering from historical medical records and employing an array of machine learning classifiers, we showcased the efficacy of eXtreme Gradient Boosting (XGBoost) model, in predicting 30-day mortality using EHR trajectory features,[35].

## 3. METHODS

### 3.1. Data

EHRs provide a longitudinal perspective of patients’ interactions with hospital service. In Norway, with predominantly public specialist healthcare, patients often have long and continuous histories within one hospital’s records. This study harnessed EHRs from St. Olavs University Hospital, Trondheim, Norway, encompassing 35,591 patients with suspected bloodstream infections (BSIs) identified via physician-initiated blood cultures between 2015 and 2020. The EHRs encompassed, curated data from the inception of electronic records in 1999 until 2020, exclusively included hospital care episodes (excluding primary care and other specialist care episodes), Intensive Care Unit (ICU) admission details, microbiology test results, laboratory test results and patient demographics comprising of gender, date of birth, and date of death. Diagnoses and Procedures within these records were classified using the International Classification of Diseases, 10th Revision (ICD-10), facilitating standardized disease identification critical for the analytical models. This study adhered to the ‘transparent reporting of a multivariable prediction model for individual prognosis or diagnosis (TRIPOD),[36]. For ethical considerations the EHRs were de-identified and accessed through a private cloud computing platform.

### 3.2. XRAI framework

The XRAI framework aggregated various data types from the raw EHRs including demographics, laboratory tests including microbiology tests, discharge summaries, and ICU admissions, as depicted in Figure 1. This dataset underwent preprocessing, event log creation, and feature engineering before being transformed and scaled to facilitate model development.

**Figure 1.**
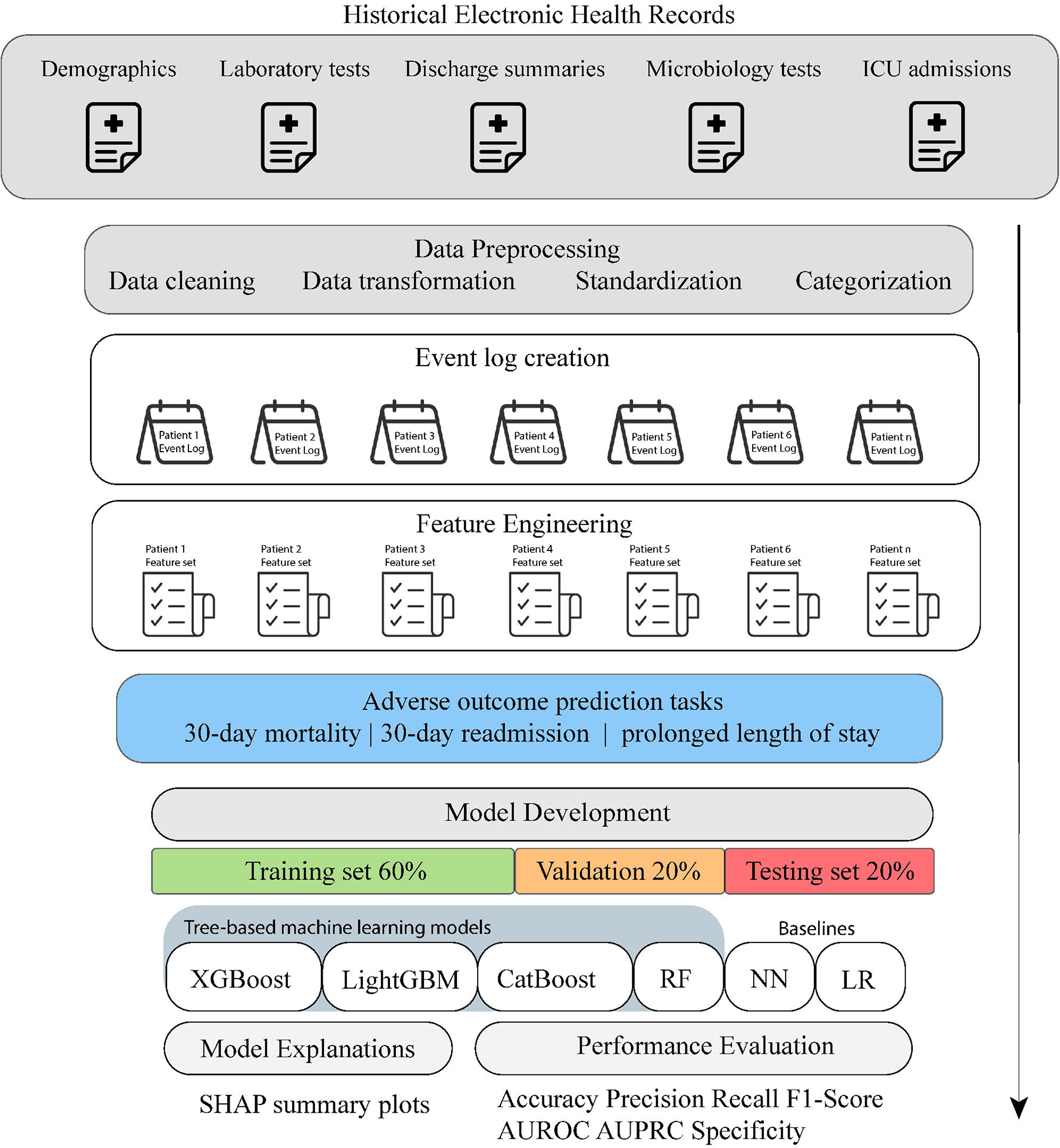
The architecture of the XRAI framework. presents the architecture of the XRAI framework. The data input sources include demographics, laboratory tests, discharge summaries, microbiology tests, and ICU admissions. This information undergoes preprocessing steps such as event log creation, feature engineering, and data transformation and scaling, to prepare it for the subsequent model development phase. The model development process involves partitioning the data into training (60%), validation (20%), and testing sets (20%) and applying tree-based machine learning algorithms XGBoost, LightGBM, CatBoost, and RF alongwith traditional NNs and LR. Performance metrics assessed included accuracy, precision, recall, F1-score, AUROC, and AUPRC. Additionally, the framework provides model explanations through SHAP summary plots.

#### 3.2.1. Data preprocessing

The medical data, initially stored in a Postgres database, were converted into CSV files to facilitate easier manipulation and access. Utilizing Python libraries such as Pandas and NumPy, the CSV files were loaded into dataframes for further processing. The episode discharge summaries required several data cleaning steps to ensure quality and relevance of the data: Relevant patient information such as identifiers, admission and discharge times, and diagnostic codes were retained. Instances of missing identifiers were addressed by replacing empty strings with NaN values and subsequently removing these records. Data were organized by patient identifier and admission/discharge times to maintain coherent episode tracking. Non-standard characters within diagnostic codes, such as semicolons and commas, were standardized to spaces, and any duplicates were removed. The timestamps were converted into datetime format, facilitating the calculation of the length of stay in hours for each episode. The duration of each ICU stay was calculated in hours, along with the total count of each type of hospital admissions per patient. The request dates in both laboratory test results and microbiology test results were standardized to datetime objects and used to create eventlogs of tests per patient. Specialized functions were created to clean the laboratory an microbiology test table entries. This function performed tasks to remove any non-numeric characters, which could represent encoding errors or artifacts from data entry. It standardized decimal point characters by replacing commas with periods, which is necessary for consistent numerical representation across different regions that may use varying formats for decimal points. The microbiology tests results column was processed to standardize and clean the values, categorizing them as ‘positive’, ‘negative’, or ‘contaminant’ based on the list of contaminants given in Supplementary Table 9. For each patient ID, four event logs were created, from episode discharge summaries, ICU admissions, laboratory tests and microbiology tests, where hospital visit/admissions, ICU admissions, particular laboratory and microbiology tests were considered as separate and overlapping medical events respectively.

#### 3.2.2. Event log creation

*Discharge Summaries Event Log:* Captures patient discharge information, including admission and discharge times, diagnostic and procedural codes, urgency and care level code.

*ICU Admissions Event Log:* Records details of each ICU stay, including the duration in hours and the total count of ICU admissions per patient and total length of ICU stays per patient.

*Laboratory Tests Event Log:* Includes results of various laboratory tests standardized and organized chronologically for each patient.

*Microbiology Tests Event Log:* Consists of microbiology test results, grouped by collection sample type, and categorizes them based on outcomes such as ‘positive’, ‘negative’, or ‘contaminant’.

#### 3.2.3. Feature Engineering

This approach involved enhancing the dataset with additional attributes derived from diagnostic and procedure codes, laboratory, and microbiology test results. From the discharge summaries predictors included, counts of the occurrences of different ICD and procedure codes, which were classified according to the initial character (alphabetic), corresponding to the various chapters of the ICD-10. Each patient record was expanded with new columns corresponding to each character, incrementing the count for each instance where a character led the code. Further stratification was conducted by categorizing the ICD-10 codes into disease groups pertinent to clinical significance, such as ‘explicit sepsis’, ‘infection’, and ‘organ dysfunction’, among others. The table depicting the ICD-10 codes selected for different disease groups is given in (Supplementary Table 8). Laboratory test results were organized using pivot tables, ensuring a structured format for analysis. Tests such as ‘Bilirubin’ (total, conjugated, and unconjugated), ‘C-Reactive Protein’ (CRP), and ‘Lactate’ (various measurements) were included, alongside white blood cell count (leukocytes), platelet count (thrombocytes), and blood gas measurements (pH, PO2). Similarly, microbiology test results were consolidated to reflect various sample types such as blood, urine, and other fluids, employing a dictionary mapping to streamline similar types. The resulting pivot table included columns for diverse samples, ranging from ‘blood culture test’ to ‘urine’, ‘feces’, and ‘nasal swabs’. The groups of microbiology tests based on sample type is given in Supplementary List Comorbidities were extracted and processed to identify unique diseases from patient records. New columns were created for each disease, and the counts were updated to represent total number of previous disease episodes in the patient records. Finally, aggregate columns were added for both ICD and procedure codes to calculate cumulative sums leading upto an episode. This method enabled the capturing of the cumulative history of medical conditions and procedures for each patient. All the NaN values representing absence of a condition or measurement were filled with zero and “0” is not interpreted as a value.

#### 3.2.3. Model Development

The development phase employed tree-based ML algorithms eXtreme Gradient Boosting (XGBoost), Light Gradient Boosting Machine (LightGBM), Category Boosting (CatBoost), and Random Forest (RF) and constructed baseline models using Neural Networks (NN) and Logistic Regression (LR) for comparative analysis. Tree-based ML models were selected primarily for their ability to handle tabular data effectively, owing to their inherent characteristics such as insensitivity to feature scaling and rotational variance, and robustness against irrelevant features (refer to Supplementary Methods for model selection rationale). Model performance was assessed using various metrics including Accuracy, Precision, Recall (Sensitivity), F1-score, Specificity, Area Under the Precision-Recall Curve (AUPRC), and Area Under the Receiver Operating Characteristic (AUROC), with definitions for each metric detailed in Supplementary Methods. For model interpretation, the framework integrated SHapley Additive exPlanations (SHAP) to attribute model outputs to individual features, leveraging a game-theoretic approach to ensure equitable feature importance distribution [23]. SHAP’s suitability for tree-based models was enhanced by its efficient computation of exact SHAP values using the TreeExplainer algorithm [37], which significantly reduced computational complexity by exploiting the structural properties of decision trees [38].

### 3.3. Predicted Outcomes

Our study aimed to predict four critical outcomes using the XRAI framework: the risk of 30-day mortality at the time of discharge, the risk of 30-day mortality at the time of admission, the risk of readmission within 30 days following discharge, and the risk of PLOS (>2 days) at the time of admission. The index episode was the last recorded hospital episode of all the patients for the mortality and PLOS prediction tasks and second last recorded episode of all patients for the readmission prediction task. We observed the following outcomes among the 35,591 patients included in the analysis: For 30-day Mortality Class 0 (no mortality within 30 days) comprised 28,173 patients (79.2%), while Class 1 (mortality within 30 days) included 7,418 patients (20.8%). For 30-day readmission following discharge, Class 0 (no readmission) was represented by 29,655 patients (83.3%), and Class 1 (readmission) had 5,936 patients (16.7%). For PLOS the Class 0 (no PLOS) consisted of 26,737 patients (75.1%), whereas Class 1 (PLOS) included 8,854 patients (24.9%). The flowchart in Figure 2 illustrates the patient distribution across these outcomes.

**Figure 2.**
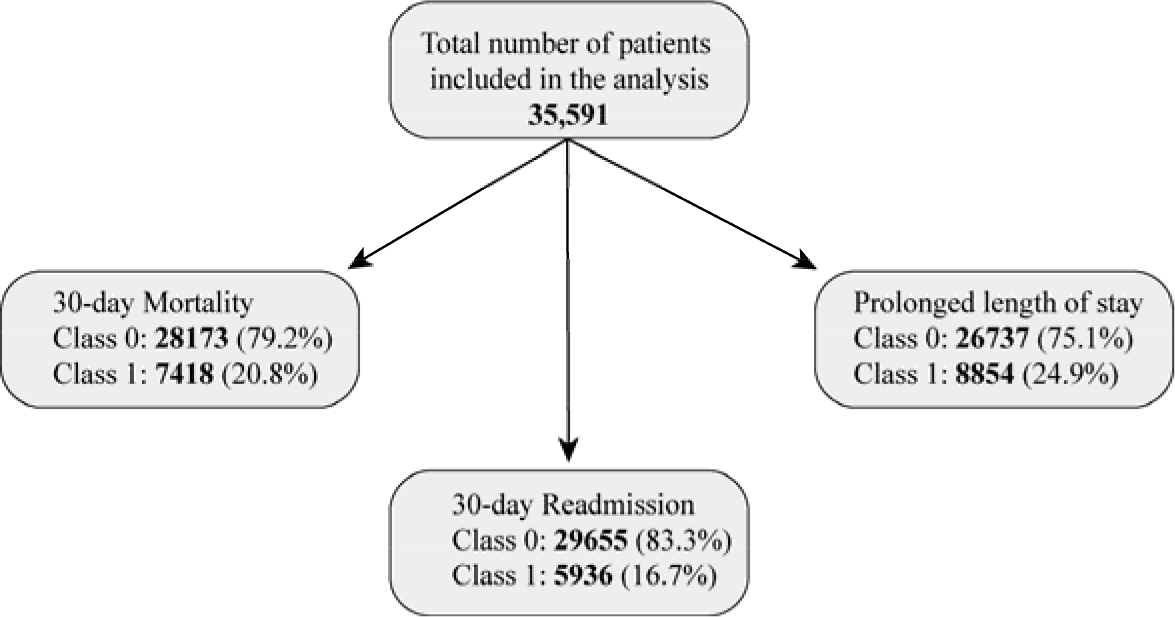
Flowchart depicting the distribution of patients for various prediction tasks.

### 3.4. Predictors

The predictors included age, sex, results of recent laboratory tests, counts of comorbidities, count of diagnostic and procedural codes, current, recent, and total hospital and ICU LOS. The most common laboratory tests were bilirubin, C-reactive protein (CRP), creatinine, leukocytes, and thrombocytes. The counts of prior positive results of microbiology tests grouped by their collected sample type were calculated and used as indicators of previous history of infections. The prediction task modelling details are given in Supplementary Methods Section 1.4.

### 3.5. Statistical Analysis of predictors

To investigate the strength and significance of predictors across the predicted outcomes in our study, we employed statistical analysis, utilizing both non-parametric and categorical data analysis techniques. For numerical features, the Mann-Whitney U test was applied to compare distributions between two independent groups defined by the outcome variables. Each feature’s mean value for both outcome classes was calculated to quantify the average influence of the feature within each group. For categorical features, such as gender, care level code, and urgency code, we conducted chi-square tests of independence.

### 3.6. Ethics Statement

This study utilizes a de-identified dataset comprising EHRs from the St. Olavs University Hospital, Trondheim, Norway, Warehouse services. The data was accessed and analysed through a secure private cloud platform. The use of the EHRs in this project has been approved by the Regional Commitees for Medical and Health Research Ethics (REK) in Central Norway by REK no. 2020/26184.

## 4. RESULTS

### 4.1. Model Evaluation

The table 1 depicts the performance metrics for 30-day mortality prediction at discharge and at admission. For the prediction of 30-day mortality at the end of the episode, the XGBoost, LightGBM, and Catboost model demonstrated superior performance with an AUROCs ranging from (0.9600-0.9625). The XGBoost model achieved the highest AUPRC and F1 score of 0.8575 and 0.7552. Similarly, For predicting 30-day mortality at the start of the episode, the tree-based models AUROCs ranged from (0.9515-0.9545). The LightGBM model achieved an AUPRC of 0.8419 and recall of 0.7324.

**Table 1.** Model performance metrics for the mortality prediction tasks.

| Models | Accuracy | Precision | Recall | F1 score | Specificity | AUPRC | AUROC (95% CI) |
| --- | --- | --- | --- | --- | --- | --- | --- |
| *30-day mortality prediction at discharge* |  |  |  |  |  |  |  |
| XGBoost | 0.9130 | 0.8221 | 0.7478 | 0.7832 | 0.9570 | 0.8524 | **0.9600 (0.9553 - 0.9642)** |
| LightGBM | 0.9168 | 0.8332 | 0.7552 | 0.7923 | 0.9598 | 0.8575 | **0.9625 (0.9580 - 0.9666)** |
| Catboost | 0.9177 | 0.8399 | 0.7512 | 0.7931 | 0.9619 | 0.8566 | **0.9625 (0.9580 - 0.9667)** |
| ANN | 0.8911 | 0.7571 | 0.7090 | 0.7323 | 0.9395 | 0.8243 | **0.9322 (0.9251 - 0.9394)** |
| RF | 0.9056 | 0.8532 | 0.6649 | 0.7474 | 0.9696 | 0.8172 | **0.9543 (0.9493 - 0.9591)** |
| LR | 0.8320 | 0.6459 | 0.4428 | 0.5254 | 0.9355 | 0.6891 | **0.8257 (0.8133 - 0.8381)** |
| *30-day mortality prediction at admission* |  |  |  |  |  |  |  |
| XGBoost | 0.9004 | 0.7885 | 0.7184 | 0.7518 | 0.9488 | 0.8336 | **0.9515 (0.9464 - 0.9564)** |
| LightGBM | 0.9055 | 0.8004 | 0.7324 | 0.7649 | 0.9515 | 0.8419 | **0.9540 (0.9493 - 0.9588)** |
| Catboost | 0.9059 | 0.8031 | 0.7311 | 0.7654 | 0.9523 | 0.8417 | **0.9545 (0.9494 - 0.9592)** |
| ANN | 0.9592 | 0.6794 | 0.6776 | 0.6785 | 0.9150 | 0.7963 | **0.9099 (0.9022 - 0.9181)** |
| RF | 0.8893 | 0.8126 | 0.6147 | 0.6999 | 0.9623 | 0.7885 | **0.9459 (0.9403 - 0.9513)** |
| LR | 0.8137 | 0.5865 | 0.3833 | 0.4636 | 0.9282 | 0.6557 | **0.7901 (0.7765 - 0.8029)** |

The table 2 depicts the performance metrics for the readmission and PLOS prediction tasks. The task of predicting readmission risk saw the CatBoost model reaching an AUROC of 0.8198, accompanied by a 95% CI of 0.8064 to 0.8325, but F1-score and Recall was low for all the models. Lastly, for the prediction of PLOS at the start of the episode, the LightGBM model proved to be highly effective, with an AUROC of 0.9797 and AUPRC of 0.9365. In all the prediction tasks XGBoost, LightGBM, and CatBoost performed better than the traditional models like ANN, RF, and LR, with LR giving the overall worst performance.

**Table 2.** Model performance metrics for the readmission and prolonged length of stay prediction tasks.

| Models | Accuracy | Precision | Recall | F1 score | Specificity | AUPRC | AUROC (95% CI) |
| --- | --- | --- | --- | --- | --- | --- | --- |
| *30-day readmission prediction at discharge* |  |  |  |  |  |  |  |
| XGBoost | 0.8460 | 0.5867 | 0.2803 | 0.3794 | 0.9602 | 0.6202 | **0.8001 (0.7867 - 0.8133)** |
| LightGBM | 0.8525 | 0.6593 | 0.2510 | 0.3636 | 0.9738 | 0.6124 | **0.8118 (0.7982 - 0.8245)** |
| Catboost | 0.8563 | 0.6886 | 0.2628 | 0.3804 | 0.9760 | 0.6194 | **0.8198 (0.8064 - 0.8325)** |
| ANN | 0.8038 | 0.4029 | 0.3506 | 0.3749 | 0.8952 | 0.6229 | **0.6839 (0.6660 - 0.7024)** |
| RF | 0.8449 | 0.7333 | 0.1197 | 0.2058 | 0.9912 | 0.5554 | **0.8009 (0.7864 - 0.8143)** |
| LR | 0.8278 | 0.4124 | 0.0611 | 0.1064 | 0.9824 | 0.5218 | **0.6141 (0.5968 - 0.6294)** |
| *PLOS prediction at admission* |  |  |  |  |  |  |  |
| XGBoost | 0.9286 | 0.8200 | 0.9179 | 0.8662 | 0.9322 | 0.9251 | **0.9779 (0.9750 - 0.9809)** |
| LightGBM | 0.9313 | 0.8115 | 0.9470 | 0.8740 | 0.9261 | 0.9365 | **0.9797 (0.9770 - 0.9822)** |
| Catboost | 0.9310 | 0.8137 | 0.9414 | 0.8729 | 0.9276 | 0.9345 | **0.9794 (0.9767 - 0.9821)** |
| ANN | 0.8941 | 0.7783 | 0.8096 | 0.7937 | 0.9225 | 0.8660 | **0.9551 (0.9504 - 0.9593)** |
| RF | 0.9237 | 0.8050 | 0.9196 | 0.8585 | 0.9251 | 0.9224 | **0.9733 (0.9702 - 0.9764)** |
| LR | 0.7665 | 0.5931 | 0.2295 | 0.3309 | 0.9471 | 0.5883 | **0.7388 (0.7251 - 0.7513)** |

### 4.2. Model Explanations for the Four Prediction Tasks

The Figure 3 depicts the SHAP summary plots for 30-day mortality prediction at discharge and at admission. For the 30-day mortality prediction at the end of the episode, important features like age, total length of stay (total_los), CRP levels, and time to the most recent hospital admission (time_to_last) stand out. High values of CRP and older age are shown to increase the risk of mortality, as indicated by the accumulation of red dots on the right side of the zero line. Similarly, in predicting 30-day mortality at the start of the episode, age and total length of stay are prominent, and in predicting 30-day mortality at the end of the episode, age and time to the most recent hospital admission are prominent. The SHAP summary plot highlights the consistent significance of these features across the different stages of hospitalization. Additionally, care level codes and the urgency of the case are influential, suggesting that more acute presentations are associated with higher mortality risk.

**Figure 3.**
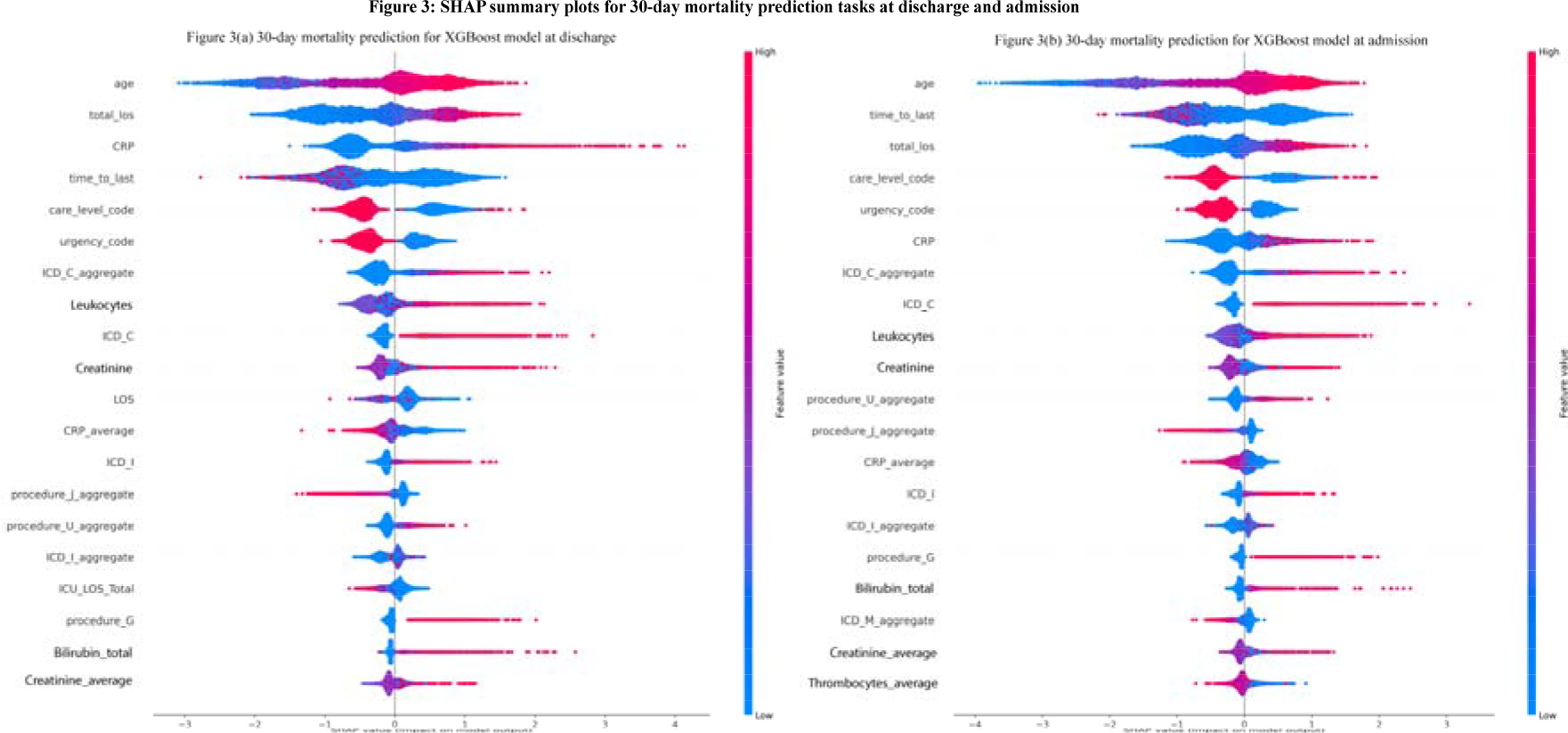
The SHAP summary plots for 30 day mortality prediction at discharge (right side) and at admission (left side) of the episode. Beeswarm plot detailing the individual SHAP values for each feature and their impact on the model’s output.

The figure 4 depicts the SHAP summary plots for the readmision risk at discharge and PLOS risk at admission. The readmission risk prediction at the end of the episode emphasizes features like time to the most recent hospital admission, age, urgency code, LOS, CRP levels, leukocytes count, and previous ICD codes related to infections (ICD_C). For the prediction of prolonged length of stay (PLOS) at the start of the episode, features like care level code, total LOS (total_los), ICD codes of ICD chapter X (J00-J99) concerning with Diseases of the respiratory system, CRP levels, and ICU length of stay (total_ICU_LOS) provide substantial predictive power. The impact of higher care levels and previous high CRP levels indicates a potentially more complicated hospital course, leading to longer stays.

**Figure 4.**
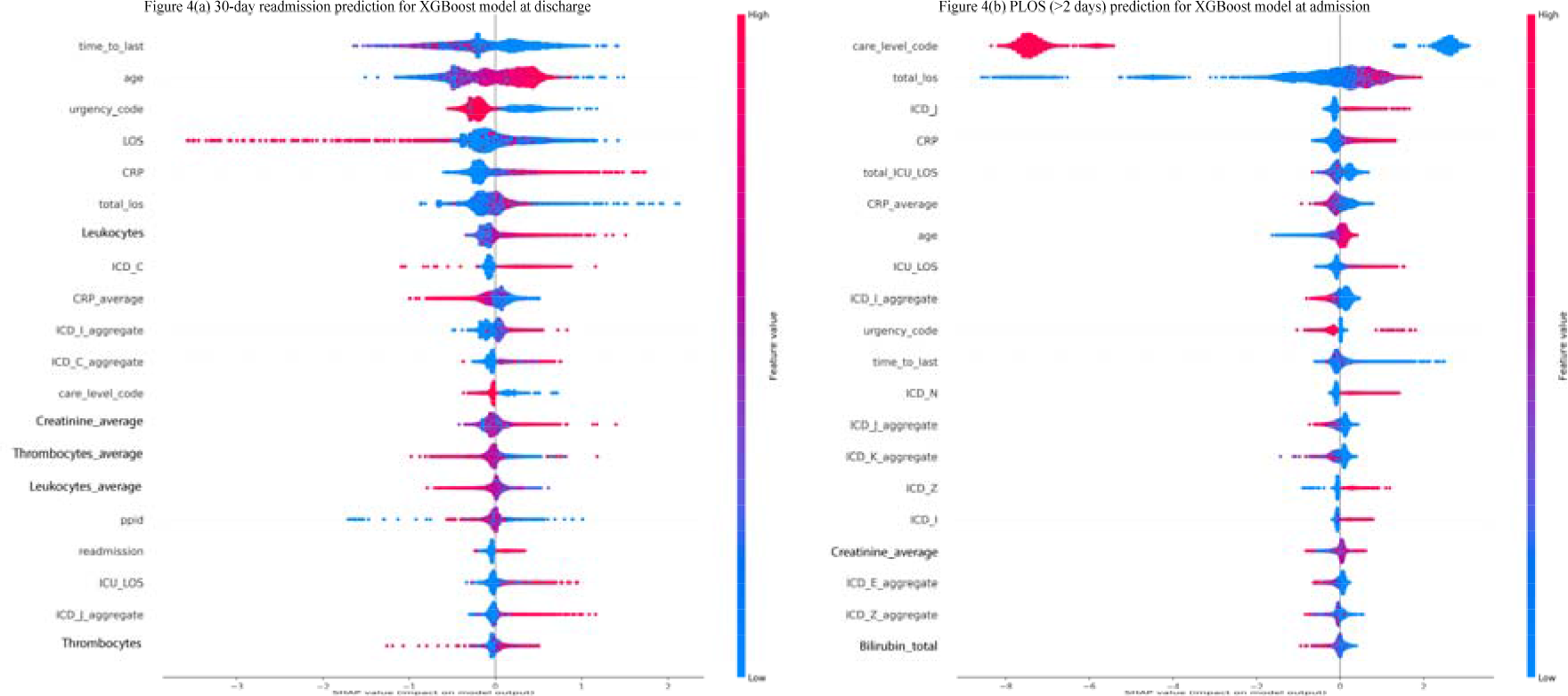
The SHAP summary plots for readmission risk at discharge (left) and PLOS risk at admission. Beeswarm plot detailing the individual SHAP values for each feature and their impact on the model’s output.

### 4.3. Patient characteristics and results of statistical analysis

The dataset’s mean patient age was 63.6 years, with a near-equal gender distribution (47.4% female, 52.5% male). The statistical analysis are detailed in Supplementary Tables 3-7. Notably, age, total length of stay, time to the recent episodes, CRP levels, urgency code, and care level code were identified as significant numerical predictors with a profound impact on the risk assessments for 30-day mortality, readmission, and PLOS. The Supplementary Table 3 gives the description for each feature and the mean values across the dataset prepared for 30-day mortality prediction at the end of the episode task. The distribution of predictors across different outcome scenarios provided insightful revelations. For instance, the statistical analysis underscored a strong association between higher CRP levels, advanced age, and an increased risk of 30-day mortality and PLOS, which aligns with clinical expectations. Similarly, the analysis of categorical predictors, such as urgency code and care level codes, unveiled significant associations with the outcomes, thus providing a deeper understanding of the factors influencing patient risk profiles.

## 5. DISCUSSIONS

Main findings of this study highlight the predictive capacity of medical history features for mortality and PLOS. Across all the four prediction tasks the tree-based models have consistently outperformed traditional ML models in efficiency, underscoring the superiority of tree-based models on tabular data,[39]. The performance for predicting mortality saw a marginal decrease from the episode’s conclusion to at the start of the episode, underscoring the important role of current episode information in forecasting mortality outcomes. Moreover, readmission prediction results were less impressive when compared with the other three tasks. Which aligns with the broader literature, suggesting a lack of strong association between hospital readmission rates and mortality across various conditions, thereby affirming the distinct pathways influencing mortality and readmission outcomes,[40, 41]. Our findings on simultaneous good results for mortality and PLOS prediction tasks are consistent with the findings on positive correlations between these two events at patient and hospital levels,[42].

In this study, we presented an comprehensive, and accurate data modelling methodology compared to modelling techniques like, temporal sequence modelling and patient data simulations, which may introduce unnecessary bias in the original data by the use of imputations,[43]. Our methodology provides a more detailed analysis at the individual patient level by including the entire medical history in the analysis, compared to similar studies which have made significant contributions to this field,[17, 18, 34]. Cai et al. developed a Bayesian Network model that, while impressive, achieved slightly lower accuracy and AUROC values compared to our models for mortality prediction,[17]. They provided a daily prediction using features from recent and current medical episodes for prediction of outcomes of the next episode, in contrast we included all the synthesized information from complete medical history available to predict impending adverse outcomes. While Tavakolian et al.’s Genetic Algorithm-Optimized Convolutional Neural Network (GAOCNN) approach to predicting hospital readmission and LoS using specific disease population datasets achieved high accuracies,[18], our study’s emphasis on ease of adoption and explainability provides more foundational value for clinical decision-making processes. The multistate model by Clark et al. for surgical patients offers a comprehensive view of hospital quality of care,[34]. Our framework complements such models by harnessing predictors of adverse hospital outcomes from the medical history for general hospital population. The limitation of this study is its reliance on data from a single center, which may not represent the diverse patient demographics hence, prevalence rates and performance metrics cannot be generalizable. Our methodology is to some extent generalizable and reproduceable. By creating medical event logs from administrative records, followed by feature sets representation of patient per prediction task, tree-based ML models, our presented framework can be implemented in all the hospitals with improved early warning systems at marginal computing costs and expertise.

## 6. CONCLUSIONS

This study presents a simple and intuitive XAI framework which comprehensively captures the complete medical history of a patient to accurately predict the risk of hospital adverse outcomes. The XRAI framework is the first of its kind to significantly enhances the predictive analysis by integrating information stored as diagnostic and procedural ICD-10 codes and deriving novel temporal features capturing critical indicators of individual trajectories. We also demonstrate that tree-based ML models, particularly XGBoost, CatBoost and LightGBM, excel in predicting these critical healthcare outcomes. The SHAP values for model explanation provide valuable insights into the decision-making process of the framework. Our findings underscore the importance of age, total length of hospital stay, recent CRP levels, care level codes, and time to the most recent hospital admission as significant predictors of patient outcomes. To further enhance the generalizability of our findings and adoption of our framework, there is a need to validate our framework on administrative datasets of hospitals outside Norway and include more diverse data sources, such as imaging data, genomics, and patient-reported outcomes in future works.

## Data Availability

All data produced in the present study are available upon reasonable request to the authors

https://github.com/EngineerRajeev/Hospital_adverse_outcomes_prediction.git

## Acknowledgements

We would like to thank the researchers at Mid-Norway Centre for Sepsis Research for valuable discussions and feedback.

## Author’s contributions

RB and Ø N conceptualized and designed the study, with input from LGT, JEA, BE and JKD. RB performed data analysis and framework development. RB wrote the initial draft of the paper, to which the rest of the authors provided comments. All authors reviewed and approved the final manuscript.

## Availability of data and materials

Data and materials are available on reasonable request to the corresponding author.

## Ethics

The use of the EHRs data in this project has been approved by the Regional Commitees for Medical and Health Research Ethics (REK) in Central Norway by REK no. 2020/26184.

## Conflict of interests

None declared.

## Funding

Financial support for this study was provided by the Computational Sepsis Mining and Modelling project through the Norwegian University of Science and Technology Health Strategic Area.

## Notes

### Competing Interest Statement

The authors have declared no competing interest.

