## Supplementary Information for "Inhospital Mortality, Readmission, and Prolonged Length of Stay Risk Prediction Leveraging Historical Electronic Health Records"

### 1. SUPPLEMENTARY METHODS

#### 1.1. ML Models:

1.1.1. LightGBM Model: The LightGBM model, a gradient boosting framework, is utilized with the scale_pos_weight parameter adjusted to address class imbalances. This model was selected for its efficiency and speed in handling large-scale data. Its gradient boosting framework is capable of processing the extensive dataset with remarkable accuracy and minimal computational resources. LightGBM's use of histogram-based decision trees allows for faster training speed and lower memory usage, making it highly suitable for the complex task of BSI prediction.

LightGBM builds the model in a stage-wise fashion and generalize them by optimizing a loss function:

$$\hat{y}(x) = \sum_{k=1}^{K} f_k\,(x)$$

Where $f_k$ are the individual decision trees and $k$ is the number of boosting stages (trees).

1.1.2. CatBoost Model: CatBoost is another gradient boosting model that is particularly effective in processing categorical features. Tuned for binary classification, it also focuses on handling class imbalance, a critical aspect in predicting BSIs.

CatBoost builds the model in a stage-wise fashion and generalize them by optimizing a loss function:

$$\hat{y}(x) = \sum_{k=1}^{K} f_k\,(x)$$

Where $f_k$ are the individual decision trees and $k$ is the number of boosting stages (trees).

1.1.3. XGBoost Model: The XGBoost (Extreme Gradient Boosting) model, stands out for its advanced regularization features is employed with a focus on performance and speed. It uses the scale_pos_weight parameter for class imbalance and a logloss evaluation metric, making it suitable for binary classification.

XGBoost builds the model in a stage-wise fashion and generalize them by optimizing a loss function:

$$\hat{y}(x) = \sum_{k=1}^{K} f_k\,(x)$$

Where $f_k$ are the individual decision trees and $k$ is the number of boosting stages (trees).

1.1.4. ANN Model: A simple Artificial Neural Network (ANN) with three dense layers (128, 64, and 1 unit) is used, featuring ReLU activation for the first two layers and sigmoid activation for the output layer. This structure allows for modeling complex non-linear relationships in the data. The model is compiled using the Adam optimizer and binary cross-entropy loss. The ANN defined by the following equations, layer-wise:

$$z^{[l+1]} = W^{[l]} a^{[l]} + b^{[l]}$$
$$a^{[l+1]} = g^{[l]} (z^{[l+1]})$$

Where $W^{[l]}$ and $b^{[l]}$ are the weights and biases at layer $l$, $a^{[l]}$ is the activation from the previous layer, $g^{[l]}$ is the activation function, and $z^{[l+1]}$ is the linear combination input to the activation function at layer $l + 1$.

1.1.5. RF Model: The Random Forest (RF) classifier with 100 estimators is used for its robustness and effectiveness in handling high-dimensional data, making it suitable for complex medical datasets. A Random Forest aggregates the predictions of multiple decision trees, typically constructed with some form of randomness and then averaged to improve the predictive accuracy and control over-fitting.
The prediction for a new sample $x$ is:

$$\hat{y} = \frac{1}{N} \sum_{i=1}^{N} t_i (x)$$

Where $t_i$ are the individual decision trees and $N$ is the number of trees in the forest.

1.1.6. LR Model: A Logistic Regression (LR) model is employed for binary classification, known for its simplicity and interpretability, which is crucial in medical settings where understanding model decisions is essential. The logistic function can be represented mathematically as:

$$P(y = 1|x) = \sigma(W^T x + b)$$

Where $\sigma (z) = \frac{1}{1+e^{-z}}$ is the logistic (sigmoid) function, $W$ is the weight vector, $x$ is the input feature vector, and b is the bias.

### 1.2. Performance Metrics:

#### 1.2.1. Accuracy:

This is the ratio of correctly predicted observations (both true positives and true negatives) to the total observations. It's a general indicator of a model's performance.

$$Accuracy = \frac{TP + TN}{TP + FP + FN + TN}$$

Where TP = True Positives, TN = True Negatives, FP = False Positives, FN = False Negatives.

#### 1.2.2. Precision:

Also called Positive Predictive Value, it is the ratio of correctly predicted positive observations to the total predicted positive observations. It shows the model's ability to return relevant results.

$$Precision = \frac{TP}{TP + FP}$$

#### 1.2.3. Recall (Sensitivity or True Positive Rate):

This is the ratio of correctly predicted positive observations to all actual positives. It measures the model's capability to find all relevant cases.

$$Recall = \frac{TP}{TP + FN}$$

#### 1.2.4. F1-score:

The F1-score is the harmonic mean of precision and recall, providing a balance between the two metrics. It's especially useful when the class distribution is uneven.

$$F1 - score = 2 \times \frac{Precision \times Recall}{Precision + Recall}$$

#### 1.2.5. Specificity (True Negative Rate):

This is the ratio of correctly predicted negative observations to all actual negatives. It measures the model's ability to identify negative results.

$$Specificity = \frac{TN}{TN + FP}$$

#### 1.2.6. Area Under the Precision-Recall (PR) Curve (AUPRC):

This metric summarizes the trade-off between the true positive rate (recall) and the positive predictive value (precision) for a predictive model using different probability thresholds. The AUPRC is particularly insightful in the case of imbalanced datasets. The PR curve plots precision (y-axis) and recall (x-axis) for different threshold values, and AUPRC is the area under this curve.

#### 1.2.7. Area Under the Receiver Operating Characteristic (AUROC):

This is used to evaluate the performance of a binary classification system by plotting the true positive rate (recall) against the false positive rate (1 - specificity) at various threshold settings. The AUROC is the area under the ROC curve, which ranges from 0 to 1. A model that predicts perfectly has an AUROC of 1, while a model that predicts randomly has an AUROC of 0.5. The ROC curve plots sensitivity (y-axis) versus 1-specificity (x-axis) for different threshold values, and AUROC is the area under this curve.

### 1.3. Training Procedure

The training of the ML models followed a structured and systematic process to ensure robustness and generalizability. The procedure incorporated several key steps.

#### 1.3.1 Data Splitting:

The initial step involved splitting the dataset into training and testing subsets to validate the model's performance. We used the **train_test_split** method from the **sklearn.model_selection** library, setting aside 20% of the data for testing. This split was performed without shuffling to preserve the temporal nature of the data. Further, the training data was divided into a smaller training set and a validation set, with 20% of the training data allocated for validation. This additional split allowed for the tuning of hyperparameters and the assessment of the model's performance during training.

#### 1.3.2. Data Scaling:

Given the sequential and tabular nature of the data, the necessary transformation steps were employed to prepare it for the corresponding learning algorithms. The data was reshaped and scaled using the StandardScaler from sklearn.preprocessing. This normalization step is crucial for models that are sensitive to the scale of input features. After scaling, the data was reshaped back to its original form, ensuring compatibility with the machine learning models used.

#### 1.4. Prediction modeling

For each predicted adverse outcome. We constructed a dataset $X^*$ from the event logs using the feature engineering steps. A positive label indicated that patients had adverse outcome at the index episode, which is denoted as $X^+ \in X^*$ . Negative labels indicated that the patient did not have a adverse outcome at the index episode, which is denoted as $X^- \in X^*$. During training phase, and validation phase, we used the data $X^*$ that contains both $X^+$ and $X^-$ to train our model Z. For testing, we applied the trained models to predict patient's risk of adverse outcome.

### 2. Supplementary Results

For comprehensive details on the ICD-10 codes used in this study, please refer to the Norwegian Directorate of eHealth's ICD-10 code directory: https://finnkode.ehelse.no/#icd10/0/0/0/-1

Additional information on clinical procedures relevant to our methodology can be found at the Norwegian Clinical Procedure Code Directory (NKPK): https://www.ehelse.no/kodeverk-og-terminologi/Norsk-klinisk-prosedyrekodeverk-(NKPK)

**Supplementary Table 3. The mean values and features description for the 30-Day mortality prediction at the end of the episode model.**

| Feature Name | Mean Value | Feature Description |
|---|---|---|
| **30_day_mortality** | 0.20842347784552273 | Target Label |
| **ICU_LOS_Total** | 2.2538141665027727 | Total cumulative ICU Length of Stay (LOS) |
| **ICU_LOS** | 0.8912283536100077 | Recent/Current ICU Length of Stay (LOS) |
| **BILIRUBIN KONJUGERT_average** | 0.8625560090776018 | Bilirubin Laboratory tests results overall average |
| **BILIRUBIN TOTAL_average** | 14.002016721055439 | Bilirubin Laboratory tests results overall average |
| **BILIRUBIN UKONJUGERT_average** | 0.6217803838649 | Bilirubin Laboratory tests results overall average |
| **CRP_average** | 58.17087332099869 | CRP Laboratory tests results overall average |
| **CRP-HØYSENSITIV_average** | 1.8714467514757769 | CRP Laboratory tests results overall average |
| **KREATININ_average** | 76.23168455129111 | Creatinine Laboratory tests results overall average |
| **LAKTAT_average** | 0.016080609337660035 | Lactate Laboratory tests results overall average |
| **LAKTAT BLODGASS_average** | 0.07059401210589121 | Lactate Laboratory tests results overall average |
| **LAKTAT BLODGASS VENØST_average** | 0.018882549226596896 | Lactate Laboratory tests results overall average |
| **LAKTAT PNA_average** | 0.012005141749318644 | Lactate Laboratory tests results overall average |
| **LEUKOCYTTER_average** | 8.913558613838148 | Leukocytes Laboratory tests results overall average |
| **PH_average** | 0.646983380425 | PH Laboratory tests results overall average |
| **PH PNA_average** | 0.06739035149335518 | PH PNA Laboratory tests results overall average |
| **PO2_average** | 0.0005647495153269086 | PO2 Laboratory tests results overall average |
| **PO2 PNA_average** | 0.0862451181478464 | PO2 PNA Laboratory tests results overall average |
| **TROMBOCYTTER_average** | 238.58878842394267 | Thrombocytes Laboratory tests results overall average |
| **BILIRUBIN KONJUGERT** | 0.054901520047202945 | Bilirubin recent and latest Laboratory tests results |
| **BILIRUBIN TOTAL** | 4.811637773594447 | Bilirubin recent and latest Laboratory tests results |
| **BILIRUBIN UKONJUGERT** | 0.08049787867719367 | Bilirubin recent and latest Laboratory tests results |
| **CRP** | 33.75483034100382 | CRP recent and latest Laboratory tests results |
| **CRP-HØYSENSITIV** | 0.19955072911691157 | CRP recent and latest Laboratory tests results |

| | | |
|---|---|---|
| **KREATININ** | 63.662801223155654 | Creatinine recent and latest Laboratory tests results |
| **LAKTAT** | 0.0018515916945295156 | Lactate recent and latest Laboratory tests results |
| **LAKTAT BLODGASS** | 0.014015715583527676 | Lactate recent and latest Laboratory tests results |
| **LAKTAT BLODGASS VENØST** | 0.0020426512320530473 | Lactate recent and latest Laboratory tests results |
| **LAKTAT PNA** | 0.0005872271079767357 | Lactate recent and latest Laboratory tests results |
| **LEUKOCYTTER** | 6.738046790855369 | Leukocytes recent and latest Laboratory tests results |
| **PH** | 0.1958705852603184 | PH recent and latest Laboratory tests results |
| **PH PNA** | 0.003326964682082549 | PH PNA recent and latest Laboratory tests results |
| **PO2** | 0.0 | PO2 recent and latest Laboratory tests results |
| **PO2 PNA** | 0.0038998623247450195 | PO2 PNA recent and latest Laboratory tests results |
| **TROMBOCYTTER** | 118.70685941202366 | Thrombocytes recent and latest Laboratory tests results |
| **Positive_annet_test** | 0.033351128094181115 | Total number of previous positive results see (Supplementary List 1) for the groups of microbiology tests by collection sample. |
| **Positive_anus_test** | 0.022646174594700908 | Total number of previous positive results |
| **Positive_bein_test** | 0.00014048495406142003 | Total number of previous positive results |
| **Positive_biopsi_test** | 0.0018544013936107444 | Total number of previous positive results |
| **Positive_blod_test** | 0.00028096990812284006 | Total number of previous positive results |
| **Positive_blood_culture_test_test** | 0.14585147930656628 | Total number of previous positive results |
| **Positive_bronki_test** | 0.02236520468657807 | Total number of previous positive results |
| **Positive_melk_test** | 0.019386923660475962 | Total number of previous positive results |
| **Positive_edta_test** | 0.16377735944480346 | Total number of previous positive results |
| **Positive_faeces_test** | 0.04107780056755921 | Total number of previous positive results |
| **Positive_hal_test** | 0.06931527633390464 | Total number of previous positive results |
| **Positive_hud_test** | 0.025231097749431036 | Total number of previous positive results |
| **Positive_led_test** | 0.00407406366778118 | Total number of previous positive results |
| **Positive_naso_test** | 0.13879913461268298 | Total number of previous positive results |
| **Positive_plasma_test** | 0.18875558427692393 | Total number of previous positive results |
| **Positive_tunge_test** | 0.005366525245146245 | Total number of previous positive results |
| **Positive_urin_test** | 1.6970020510803292 | Total number of previous positive results |
| **Positive_ear_test** | 0.013149391700148914 | Total number of previous positive results |
| **Positive_eye_test** | 0.006799471776572729 | Total number of previous positive results |
| **urgency_code** | 2.862690005900368 | Urgency code for the current epsiode |
| **care_level_code** | 2.3205304711865358 | Care level code for the current episode |
| **LOS** | 1.983324435952872 | Length of Stay of the current episode |
| **time_to_last** | 3508.229299542019 | Time to the most recent hospital episode |
| **total_los** | 34.784812171616345 | Total cumulative hospital Length of Stay |
| **age** | 59.545755949537806 | Age at the time of prediction |
| **procedure_A** | 0.030766204939450984 | Count of preocedure codes starting with the letter |
| **procedure_B** | 0.0005619398162456801 | Count of preocedure codes starting with the letter |
| **procedure_C** | 0.02180326487033239 | Count of preocedure codes starting with the letter |
| **procedure_D** | 0.03613273018459723 | Count of preocedure codes starting with the letter |
| **procedure_E** | 0.005956562052204209 | Count of preocedure codes starting with the letter |
| **procedure_F** | 0.0481301452614425 | Count of preocedure codes starting with the letter |
| **procedure_G** | 0.08631395577533646 | Count of preocedure codes starting with the letter |

| | | |
|---|---|---|
| **procedure_H** | 0.0005057458346211121 | Count of preocedure codes starting with the letter |
| **procedure_I** | 0.0054227192267708125 | Count of preocedure codes starting with the letter |
| **procedure_J** | 0.07066393189289427 | Count of preocedure codes starting with the letter |
| **procedure_K** | 0.027057402152229498 | Count of preocedure codes starting with the letter |
| **procedure_L** | 0.008541485206934337 | Count of preocedure codes starting with the letter |
| **procedure_M** | 0.012812227810401505 | Count of preocedure codes starting with the letter |
| **procedure_N** | 0.03669467000084291 | Count of preocedure codes starting with the letter |
| **procedure_O** | 0.03315444915849512 | Count of preocedure codes starting with the letter |
| **procedure_P** | 0.01691438846899497 | Count of preocedure codes starting with the letter |
| **procedure_Q** | 0.022561883622264054 | Count of preocedure codes starting with the letter |
| **procedure_R** | 0.029726616279396476 | Count of preocedure codes starting with the letter |
| **procedure_T** | 0.008569582197746622 | Count of preocedure codes starting with the letter |
| **procedure_U** | 0.007305217611193841 | Count of preocedure codes starting with the letter |
| **procedure_W** | 0.09659745441263241 | Count of preocedure codes starting with the letter |
| **procedure_X** | 0.00022477592649827204 | Count of preocedure codes starting with the letter |
| **procedure_Y** | 0.005001264364586553 | Count of preocedure codes starting with the letter |
| **procedure_Z** | 0.034418813745047905 | Count of preocedure codes starting with the letter |
| **ICD_A** | 0.04001011491669242 | Count of Diagnostic codes starting with the letter in the current episode |
| **ICD_B** | 0.03734090078952544 | Count of Diagnostic codes starting with the letter |
| **ICD_C** | 0.2195217892163749 | Count of Diagnostic codes starting with the letter |
| **ICD_D** | 0.05178275406703942 | Count of Diagnostic codes starting with the letter |
| **ICD_E** | 0.110505464864713 | Count of Diagnostic codes starting with the letter |
| **ICD_F** | 0.058132673990615606 | Count of Diagnostic codes starting with the letter |
| **ICD_G** | 0.060380433255598324 | Count of Diagnostic codes starting with the letter |
| **ICD_H** | 0.046247646877019474 | Count of Diagnostic codes starting with the letter |
| **ICD_I** | 0.3167654744176899 | Count of Diagnostic codes starting with the letter |
| **ICD_J** | 0.21530724059453232 | Count of Diagnostic codes starting with the letter |
| **ICD_K** | 0.11179792644207805 | Count of Diagnostic codes starting with the letter |
| **ICD_L** | 0.040178696861566124 | Count of Diagnostic codes starting with the letter |
| **ICD_M** | 0.07959877497120059 | Count of Diagnostic codes starting with the letter |
| **ICD_N** | 0.14096260290522886 | Count of Diagnostic codes starting with the letter |
| **ICD_O** | 0.016099575735438736 | Count of Diagnostic codes starting with the letter |
| **ICD_P** | 0.01837543199123374 | Count of Diagnostic codes starting with the letter |
| **ICD_Q** | 0.01340226461745947 | Count of Diagnostic codes starting with the letter |
| **ICD_R** | 0.12944283667219242 | Count of Diagnostic codes starting with the letter |
| **ICD_S** | 0.04543283414346323 | Count of Diagnostic codes starting with the letter |
| **ICD_T** | 0.028096990812284003 | Count of Diagnostic codes starting with the letter |
| **ICD_U** | 0.0025849231547301283 | Count of Diagnostic codes starting with the letter |
| **ICD_V** | 0.0006462307886825321 | Count of Diagnostic codes starting with the letter |
| **ICD_W** | 0.0024163412098564246 | Count of Diagnostic codes starting with the letter |
| **ICD_X** | 0.0005900368070579641 | Count of Diagnostic codes starting with the letter |
| **ICD_Y** | 0.0037088027872214887 | Count of Diagnostic codes starting with the letter |
| **ICD_Z** | 0.21404287600797955 | Count of Diagnostic codes starting with the letter |

| | | |
|---|---|---|
| **urinarytractinfection** | 0.06821949369222556 | Count of urinary tract infection episodes in the medical history |
| **cardiovascular** | 0.2745076002360147 | Count of cardiovascular disease episodes |
| **lung** | 0.008878649096681745 | Count of lung disease episodes |
| **centralnervoussystem** | 0.000618133797870248 | Count of CNS disease episodes |
| **organdysfunction** | 0.10600994633474754 | Count of organdysfunction episodes |
| **skinandsofttissueinfection** | 0.03978533899019415 | Count of the disease episodes |
| **pneumonia** | 0.02146610098058498 | Count of the disease episodes |
| **endocarditis** | 0.001236267595740496 | Count of the disease episodes |
| **sepsis** | 0.006349919923576185 | Count of the disease episodes |
| **infection** | 0.41763367143378943 | Count of the disease episodes |
| **dementia** | 0.024865836868871345 | Count of the disease episodes |
| **explicitsepsis** | 0.0019386923660475962 | Count of the disease episodes |
| **intraabdominalinfection** | 0.0009833946784299401 | Count of the disease episodes |
| **cancer** | 0.09075328032367734 | Count of the disease episodes |
| **ICD_A_aggregate** | 0.4277485881262117 | Cumulative count in the complete history |
| **ICD_B_aggregate** | 0.5174341827990222 | Cumulative count in the complete history |
| **ICD_C_aggregate** | 9.095529768761766 | Cumulative count in the complete history |
| **ICD_D_aggregate** | 1.5142311258464218 | Cumulative count in the complete history |
| **ICD_E_aggregate** | 2.4085021494197973 | Cumulative count in the complete history |
| **ICD_F_aggregate** | 3.368520131493917 | Cumulative count in the complete history |
| **ICD_G_aggregate** | 1.597651091568093 | Cumulative count in the complete history |
| **ICD_H_aggregate** | 2.0057036891348936 | Cumulative count in the complete history |
| **ICD_I_aggregate** | 6.000814812733556 | Cumulative count in the complete history |
| **ICD_J_aggregate** | 2.5278300693995672 | Cumulative count in the complete history |
| **ICD_K_aggregate** | 2.7337529150627966 | Cumulative count in the complete history |
| **ICD_L_aggregate** | 1.6141159281840916 | Cumulative count in the complete history |
| **ICD_M_aggregate** | 3.269393947908179 | Cumulative count in the complete history |
| **ICD_N_aggregate** | 4.551909190525695 | Cumulative count in the complete history |
| **ICD_O_aggregate** | 0.39096962715293193 | Cumulative count in the complete history |
| **ICD_P_aggregate** | 0.2741704363462673 | Cumulative count in the complete history |
| **ICD_Q_aggregate** | 0.3810232924053834 | Cumulative count in the complete history |
| **ICD_R_aggregate** | 2.800455171251159 | Cumulative count in the complete history |
| **ICD_S_aggregate** | 0.9926666853979939 | Cumulative count in the complete history |
| **ICD_T_aggregate** | 0.9181253687730044 | Cumulative count in the complete history |
| **ICD_U_aggregate** | 0.033632098002303955 | Cumulative count in the complete history |
| **ICD_V_aggregate** | 0.02034222134809362 | Cumulative count in the complete history |
| **ICD_W_aggregate** | 0.04633193784945632 | Cumulative count in the complete history |
| **ICD_X_aggregate** | 0.03115956281082296 | Cumulative count in the complete history |
| **ICD_Y_aggregate** | 0.07459751060661403 | Cumulative count in the complete history |
| **ICD_Z_aggregate** | 9.554971762524234 | Cumulative count in the complete history |
| **procedure_A_aggregate** | 0.9207664859093591 | Cumulative count in the complete history |
| **procedure_B_aggregate** | 0.03087859290270012 | Cumulative count in the complete history |
| **procedure_C_aggregate** | 1.057683122137619 | Cumulative count in the complete history |

| | | |
|---|---|---|
| **procedure_D_aggregate** | 1.074485122643365 | Cumulative count in the complete history |
| **procedure_E_aggregate** | 0.10634711022449496 | Cumulative count in the complete history |
| **procedure_F_aggregate** | 1.5416818858700234 | Cumulative count in the complete history |
| **procedure_G_aggregate** | 1.4267089994661573 | Cumulative count in the complete history |
| **procedure_H_aggregate** | 0.03930769014638532 | Cumulative count in the complete history |
| **procedure_I_aggregate** | 1.5468236351886713 | Cumulative count in the complete history |
| **procedure_J_aggregate** | 1.4023208114410946 | Cumulative count in the complete history |
| **procedure_K_aggregate** | 2.251973813604563 | Cumulative count in the complete history |
| **procedure_L_aggregate** | 0.3270208760641735 | Cumulative count in the complete history |
| **procedure_M_aggregate** | 0.3729313590514456 | Cumulative count in the complete history |
| **procedure_N_aggregate** | 1.1501503189008457 | Cumulative count in the complete history |
| **procedure_O_aggregate** | 3.157876991374224 | Cumulative count in the complete history |
| **procedure_P_aggregate** | 0.3944817510044674 | Cumulative count in the complete history |
| **procedure_Q_aggregate** | 1.077070045798095 | Cumulative count in the complete history |
| **procedure_R_aggregate** | 1.3290719563934703 | Cumulative count in the complete history |
| **procedure_T_aggregate** | 0.7123430080638363 | Cumulative count in the complete history |
| **procedure_U_aggregate** | 0.8687027619341968 | Cumulative count in the complete history |
| **procedure_W_aggregate** | 4.685229411929982 | Cumulative count in the complete history |
| **procedure_X_aggregate** | 0.03627321513865865 | Cumulative count in the complete history |
| **procedure_Y_aggregate** | 0.006265628951139333 | Cumulative count in the complete history |
| **procedure_Z_aggregate** | 3.5730381276165324 | Cumulative count in the complete history |
| **prior_comorbidities_counts** | 0.9704700626562895 | Cumulative count in the complete history |
| **Gender** | 0.5246832064285915 | Cumulative count in the complete history |

**Supplementary Table 4. Sorted statistical summary table for 30-day mortality predictors at the end of the episode.**

| | Stat | P-Value | Mean (Class 0) | Mean (Class 1) | Chi2 Stat | Proportion (Class 0) | Proportion (Class 1) |
|---|---|---|---|---|---|---|---|
| **urgency_code** | | 0.0 | | | 3476.8719636480846 | 0.6887445426472154 | 0.6350768401186303 |
| **ICD_J_aggregate** | 75051088.0 | 0.0 | 2.1058460227877753 | 4.130493394445942 | | | |
| **ICD_I_aggregate** | 70409711.5 | 0.0 | 5.215170553366699 | 8.984631976273928 | | | |
| **CRP** | 54503172.5 | 0.0 | 20.847211869520457 | 82.77698391300441 | | | |
| **ICD_C_aggregate** | 66088361.5 | 0.0 | 5.717992404074823 | 21.923159881369642 | | | |
| **procedure_R_aggregate** | 80991734.0 | 0.0 | 1.203457210804671 | 1.8061472094904287 | | | |
| **procedure_0_aggregate** | 92207019.0 | 0.0 | 0.20679373868597595 | 1.8456457266109463 | | | |
| **care_level_code** | | 0.0 | | | 5309.82522571323 | 0.7020906541724345 | 0.6952008627662443 |

| | | | | |
|---|---|---|---|---|
| **LOS** | 55965554.0 | 0.0 | 1.17887250204 10022 | 5.03857171744 4051 |
| **time_to_last** | 139810218. 5 | 0.0 | 4029.56838107 4078 | 1528.22310595 84794 |
| **LEUKOCYTTER** | 66826795.0 | 0.0 | 5.59382943480 1651 | 11.0836973128 42686 |
| **ICD_I** | 78565754.5 | 0.0 | 0.20370567564 689596 | 0.74615799406 8482 |
| **ICD_C** | 71549881.0 | 0.0 | 0.10201256522 202108 | 0.66581288757 07738 |
| **age** | 55954082.5 | 0.0 | 55.4414865296 5605 | 75.1334591534 1063 |
| **total_los** | 55653342.5 | 0.0 | 29.3755131982 1564 | 55.3289183966 9267 |
| **ICD_J** | 80707904.0 | 0.0 | 0.14293827423 419586 | 0.49015907252 628743 |
| **procedure_G** | 92445021.5 | 6.4611478 08177588 e-307 | 0.05373939587 548362 | 0.21002965758 96468 |
| **KREATININ_avera ge** | 77213942.0 | 3.8725685 98236364 e-263 | 70.3545135905 4105 | 98.5527330114 1723 |
| **ICD_D_aggregate** | 81522390.0 | 1.0225342 24172198 6e-248 | 1.32261385014 02052 | 2.24197897007 2796 |
| **procedure_G_aggr egate** | 84138332.5 | 2.5276146 83077857 5e-213 | 1.26567990629 32595 | 2.03828525208 9512 |
| **ICD_E_aggregate** | 83236161.5 | 2.5274355 46142632 e-204 | 2.26273382316 4022 | 2.96211916958 74898 |
| **procedure_U_aggr egate** | 84930257.0 | 9.1179932 54886273 e-203 | 0.72927980690 73226 | 1.39822054462 11917 |
| **prior_comorbiditie s_counts** | 82260989.5 | 1.0661305 60647812 9e-197 | 0.88620310226 103 | 1.29050957131 30224 |
| **Positive_urin_test** | 83689866.5 | 3.4100969 88898291 6e-182 | 1.57004933801 86704 | 2.17915880291 18363 |
| **cancer** | 93554126.5 | 8.9758234 85430375 e-180 | 0.06680154758 101729 | 0.18172014019 951468 |
| **procedure_R** | 98428871.0 | 3.0903051 00186988 e-166 | 0.01558229510 5242608 | 0.08344567268 805608 |
| **KREATININ** | 84065630.0 | 9.4758215 74673034 e-152 | 55.1759259456 45346 | 95.8953082591 8935 |
| **procedure_P_aggr egate** | 90861981.0 | 9.0957427 6985113e-142 | 0.33454016256 699676 | 0.62213534645 45699 |
| **cardiovascular** | 89793234.5 | 1.5143070 95009835 6e-134 | 0.24228871614 666525 | 0.39687247236 45187 |

| | | | | |
|---|---|---|---|---|
| **Positive_edta_test** | 95068501.0 | 1.990367851231134e-127 | 0.1289177581372236 | 0.2961714747910488 |
| **procedure_W_aggregate** | 86694332.0 | 4.942459850275391e-125 | 3.9896354665814786 | 7.32704232946886 |
| **ICD_N** | 93989762.0 | 1.24663677316102 24e-119 | 0.11560714158946508 | 0.23726071717444056 |
| **ICD_E** | 95326940.5 | 6.173932779419837e-118 | 0.08564938061264331 | 0.20490698301428956 |
| **BILIRUBIN TOTAL** | 92208196.5 | 6.624664274016081e-117 | 3.4652740803842925 | 9.925024714658038 |
| **CRP_average** | 86630808.5 | 5.239982296305585e-114 | 55.51249149748861 | 68.26720536652978 |
| **procedure_T_aggregate** | 90388872.0 | 3.177158173687538e-113 | 0.6153764242359706 | 1.0806147209490429 |
| **ICD_N_aggregate** | 88194014.5 | 5.312688476904842e-110 | 3.7141589465090687 | 7.733620922081424 |
| **procedure_P** | 101035132.0 | 1.84268441948237 95e-108 | 0.007631420154048202 | 0.05217039633324346 |
| **procedure_Y_aggregate** | 102541509.5 | 8.605980998851064e-105 | 0.0004969296844496504 | 0.028174710164464815 |
| **procedure_Y** | 102736544.5 | 2.316684852241728e-104 | 3.549497746068931e-05 | 0.02386087894311135 |
| **Positive_blood_culture_test_test** | 95138371.0 | 7.578312641078385e-102 | 0.11745288041742094 | 0.2537071987058506 |
| **ICD_O_aggregate** | 110650265.0 | 1.56260286018624 74e-96 | 0.49135697298832215 | 0.009706120248045295 |
| **procedure_M_aggregate** | 110566746.5 | 8.144818502358003e-96 | 0.46778830795442444 | 0.012671879212725803 |
| **procedure_F_aggregate** | 90623111.5 | 7.971645053399038e-94 | 1.4430838036417848 | 1.916149905634942 |
| **ICU_LOS_Total** | 89041514.0 | 1.55794160935392 13e-89 | 1.9911159029330665 | 3.2515221982565077 |
| **ICD_A_aggregate** | 92787483.5 | 4.850371328140499e-89 | 0.38441060589926523 | 0.5923429495820977 |
| **ICD_Z** | 93957829.0 | 6.706717631485326e-82 | 0.19075000887374435 | 0.3025074143974117 |
| **Positive_hal_test** | 98708929.0 | 7.892420251186258e-77 | 0.05359741596564086 | 0.12901051496360205 |

| | | | | |
|---|---|---|---|---|
| **ICD_A** | 99770196.5 | 1.52009926562003e-71 | 0.03041919568381074 | 0.0764356969533567 |
| **Positive_plasma_test** | 96860961.0 | 6.1160736593606e-70 | 0.15855606431689917 | 0.3034510649770828 |
| **ICU_LOS** | 90988090.5 | 6.596822692764581e-70 | 0.7053310498231149 | 1.5972521793834782 |
| **ICD_D** | 99455368.0 | 5.680102566866028e-67 | 0.03911546516167962 | 0.09989215421946616 |
| **PH_average** | 110996020.5 | 1.41362397448917 43e-63 | 0.7421480177940367 | 0.28555532318614063 |
| **procedure_T** | 102468272.0 | 1.458061868973886e-62 | 0.004188407340361339 | 0.025208951199784307 |
| **ICD_P_aggregate** | 109021797.0 | 6.802837309796926e-62 | 0.3350725872289071 | 0.04286869776220006 |
| **organdysfunction** | 97949570.0 | 1.66108885744672 35e-57 | 0.09239342633017428 | 0.15772445403073604 |
| **ICD_Z_aggregate** | 92240451.5 | 1.61011202914313 34e-55 | 8.39953146629752 | 13.943246157994068 |
| **LAKTAT BLODGASS_average** | 108530444.0 | 2.13096910513560 98e-54 | 0.0844358944540068 | 0.018023595363715325 |
| **ICD_M** | 109881934.0 | 1.711999836259085e-51 | 0.09125758705143222 | 0.03531949312483149 |
| **ICD_R_aggregate** | 92940110.0 | 2.996441892051741e-51 | 2.7780854009157703 | 2.8854138581827984 |
| **ICD_H** | 108544372.5 | 1.75431041429696 75e-49 | 0.05530117488375395 | 0.011863035858722027 |
| **ICD_F** | 100167796.0 | 6.506232912619054e-47 | 0.048805594008447806 | 0.09355621461310326 |
| **Positive_bronki_test** | 101868686.0 | 2.357204198734685e-44 | 0.016150214744613638 | 0.045969263952547856 |
| **LEUKOCYTTER_average** | 93662522.5 | 4.56726134117201e-43 | 8.708393988205863 | 9.692758263061663 |
| **procedure_X_aggregate** | 101497771.5 | 3.882770842424184e-37 | 0.02903489156284386 | 0.0637638177406309 |
| **PH** | 107260758.5 | 1.63516219658533 78e-36 | 0.23660756752919415 | 0.04115462388784039 |
| **BILIRUBIN TOTAL_average** | 94705696.0 | 8.210433321430327e-36 | 14.062026597157073 | 13.774103774248708 |

| | | | | |
|---|---|---|---|---|
| **infection** | 96302059.5 | 4.3075193822821885e-35 | 0.396549888190821 | 0.49770827716365595 |
| **ICD_Q_aggregate** | 108181231.0 | 2.187826908231105e-32 | 0.4477691406665957 | 0.1275276354812618 |
| **dementia** | 102049888.0 | 1.430375398988515e-31 | 0.019912682355446704 | 0.04367754111620383 |
| **procedure_D** | 106844171.0 | 1.10248336070188704e-28 | 0.04319738756965889 | 0.009301698571043408 |
| **procedure_C** | 106377213.5 | 3.708607221870702e-28 | 0.027047172825045256 | 0.0018873011593421407 |
| **ICD_F_aggregate** | 98508343.5 | 6.300614687658106e-26 | 3.6310297092961346 | 2.371528713939067 |
| **ICD_K_aggregate** | 96903020.5 | 3.45067887158446 46e-25 | 2.7748908529443086 | 2.577514154758695 |
| **procedure_U** | 103359168.5 | 1.00061949548205 54e-23 | 0.004756326979732368 | 0.016985710434079267 |
| **procedure_O** | 106600839.5 | 1.08021490808063 25e-23 | 0.039186455116601 | 0.010245349150714478 |
| **TROMBOCYTTER_average** | 112253578.0 | 6.297300651517788e-23 | 240.82809185651757 | 230.0840842441166 |
| **Positive_naso_test** | 100421835.0 | 2.38929562418938 25e-22 | 0.1282078585880098 | 0.17902399568616878 |
| **ICD_O** | 105716330.0 | 4.987008878310634e-20 | 0.020232137152592908 | 0.0004044216770018873 |
| **procedure_L_aggregate** | 108312258.5 | 3.421784574536348e-19 | 0.3592446668796365 | 0.20463736856295497 |
| **ICD_R** | 100540188.0 | 5.245791812563757e-19 | 0.12011500372697263 | 0.16486923699110273 |
| **LAKTAT BLODGASS VENØST_average** | 105674826.5 | 2.41449693194096 24e-17 | 0.02260626942716178 | 0.004740143024047042 |
| **procedure_M** | 105483960.0 | 3.8939167 7880088e-17 | 0.016185709722074327 | 0.0 |
| **procedure_W** | 101404125.0 | 2.45332629393787 47e-16 | 0.08607532034217158 | 0.1365597196009706 |
| **procedure_Z_aggregate** | 110298205.0 | 3.58008021944778 33e-16 | 3.829872573030916 | 2.597600431383122 |
| **CRP-HØYSENSITIV** | 105749587.5 | 1.83796298140071 03e-15 | 0.14602988677102186 | 0.40281881908870315 |

| | | | | | | | |
|---|---|---|---|---|---|---|---|
| **Gender** | | 3.3766183 93448541 7e-15 | | | 62.0335116 31887876 | 0.51396727 36307812 | 0.56538150 44486384 |
| **ICD_L** | 106464572. 5 | 1.7570879 96975330 3e-14 | 0.04461718666 8086464 | 0.02332165004 0442166 | | | |
| **ICD_B_aggregate** | 100296963. 5 | 1.3652839 04706489 8e-13 | 0.51552905263 90516 | 0.52466972229 71152 | | | |
| **procedure_K_aggr egate** | 100408301. 0 | 2.1931128 26929006 7e-13 | 1.54704859262 41437 | 4.92922620652 467 | | | |
| **ICD_H_aggregate** | 100033294. 5 | 8.4408867 46405153 e-13 | 1.99275902459 80193 | 2.05486654084 65894 | | | |
| **ICD_W_aggregate** | 102595212. 0 | 1.8070273 85521336 4e-12 | 0.04152912362 9006494 | 0.06457266109 463468 | | | |
| **ICD_Y_aggregate** | 102294515. 5 | 1.0684703 91604247 8e-11 | 0.07024456039 470416 | 0.09112968455 109194 | | | |
| **ICD_M_aggregate** | 109235060. 5 | 1.5408889 04007222 2e-11 | 3.55400560820 64386 | 2.18846050148 28793 | | | |
| **sepsis** | 103765887. 0 | 1.8332224 66786690 4e-11 | 0.00489830688 9575125 | 0.01186303585 8722027 | | | |
| **procedure_L** | 105175953. 0 | 5.2344224 14085725 e-11 | 0.01047101835 0903347 | 0.00121326503 10056618 | | | |
| **TROMBOCYTTER** | 100103729. 0 | 1.2616789 33650393 7e-09 | 118.098741111 46608 | 121.016446481 53143 | | | |
| **procedure_O_aggr egate** | 108276514. 0 | 2.1972929 0443262e-09 | 3.36570475277 7482 | 2.36856295497 43865 | | | |
| **procedure_F** | 102957228. 5 | 5.1690918 03468203 e-09 | 0.04131615376 424236 | 0.07400916689 134537 | | | |
| **ICD_G** | 102650295. 0 | 6.3646433 23736832 e-09 | 0.05583359954 566429 | 0.07764896198 436236 | | | |
| **procedure_I_aggre gate** | 106982345. 5 | 1.5476062 98363324 5e-08 | 1.69683739750 82527 | 0.97708277163 65598 | | | |
| **procedure_A** | 103420001. 5 | 2.9039856 17059267 8e-08 | 0.02814751712 6326623 | 0.04071178215 1523324 | | | |
| **procedure_C_aggr egate** | 101684380. 5 | 7.1457328 28447206 e-08 | 1.05714691371 17098 | 1.05971960097 0612 | | | |
| **procedure_E** | 104999648. 0 | 7.3779203 7756632e-08 | 0.00724097540 198062 | 0.00107845780 53383662 | | | |
| **LAKTAT BLODGASS** | 105120165. 5 | 3.0673510 22151198 e-07 | 0.01458311148 9724205 | 0.01186078907 162757 | | | |

| | | | | |
|---|---|---|---|---|
| **procedure_B_aggregate** | 105417300.5 | 3.598388795057366 7e-07 | 0.03478507791 1475525 | 0.01604205985 4408198 |
| **Positive_ear_test** | 105150635.0 | 4.257154222009632 7e-07 | 0.01487239555 6028822 | 0.00660555405 7697492 |
| **procedure_J** | 106061536.5 | 5.209436631655601 e-07 | 0.07123841976 360346 | 0.06848207063 898624 |
| **ICD_G_aggregate** | 101544903.5 | 8.105848268610126 e-07 | 1.64366592127 214 | 1.42289026691 83068 |
| **explicitsepsis** | 104208325.0 | 1.385425492435936 6e-06 | 0.00138430412 09668833 | 0.00404421677 0018873 |
| **ICD_Q** | 105217534.5 | 1.776958370800763 4e-06 | 0.01448195080 396124 | 0.00930169857 1043408 |
| **procedure_0** | 104409138.0 | 1.806587735013877 2e-06 | 0.0 | 0.00080884335 40037746 |
| **procedure_E_aggregate** | 105927272.0 | 9.63951782672537e-06 | 0.11141873424 910376 | 0.08708546778 107307 |
| **ICD_U** | 104190510.5 | 1.072528293856268 e-05 | 0.00195222376 0337912 | 0.00498786734 9689944 |
| **ICD_U_aggregate** | 103843306.5 | 2.322706947488587 e-05 | 0.02541440386 185355 | 0.06484227554 596926 |
| **procedure_Q** | 105256300.0 | 3.257405883344266 e-05 | 0.02399460476 3425974 | 0.01712051765 974656 |
| **ICD_P** | 105030545.5 | 3.300800819177499 7e-05 | 0.01803144855 003017 | 0.01968185494 7425182 |
| **ICD_S_aggregate** | 101948491.0 | 9.759260890999675 e-05 | 1.00596315621 33957 | 0.94216770018 87301 |
| **procedure_A_aggregate** | 106855752.5 | 0.000107264863126 45504 | 0.93603805061 58379 | 0.86276624427 06929 |
| **BILIRUBIN UKONJUGERT_average** | 103899777.0 | 0.000601816412659 8498 | 0.64878497055 5815 | 0.51921901680 59675 |
| **ICD_V_aggregate** | 104926858.0 | 0.000681025264177 2957 | 0.01994817733 2907394 | 0.02183877055 8101916 |
| **BILIRUBIN KONJUGERT_average** | 103837037.0 | 0.000681487355829 7512 | 0.66336170277 97626 | 1.61908083939 96583 |
| **procedure_H_aggregate** | 105171323.0 | 0.000721288870429 3516 | 0.04312639761 4737515 | 0.02480452952 278242 |
| **ICD_B** | 103712851.5 | 0.001869074557896 3171 | 0.03521101764 1003796 | 0.04543003504 987867 |

| | | | | |
|---|---|---|---|---|
| **procedure_J_aggregate** | 102456777.5 | 0.00276575241155 32192 | 1.37230681858 51701 | 1.51631167430 57428 |
| **procedure_X** | 104434351.5 | 0.00371884006979 2487 | 0.00010648493 238206793 | 0.00067403612 83364788 |
| **procedure_N_aggregate** | 106353592.5 | 0.00450699207275 9957 | 1.19799098427 5725 | 0.96845510919 38528 |
| **ICD_L_aggregate** | 102845052.5 | 0.00557784235105 4158 | 1.65481134419 47965 | 1.45955783229 98113 |
| **PO2_average** | 104465484.0 | 0.00585046974121 691 | 0.0 | 0.00270962523 5912645 |
| **intraabdominalinfection** | 104605676.5 | 0.00877215850493 6182 | 0.00120682923 36634367 | 0.00013480722 566729578 |
| **Positive_faeces_test** | 103887389.5 | 0.00919980529035 4357 | 0.04089021403 471409 | 0.04179023995 686169 |
| **Positive_led_test** | 104667341.0 | 0.01121238049963 5225 | 0.00472083200 2271679 | 0.00161768670 80075492 |
| **ICD_W** | 104328414.5 | 0.01304538920818 1941 | 0.00205870869 271998 | 0.00377460231 86842814 |
| **LAKTAT PNA_average** | 104173956.5 | 0.01328729952693 1213 | 0.01111702694 0687896 | 0.01537813426 7996769 |
| **pneumonia** | 104010138.0 | 0.01430067147449 932 | 0.02051609697 2278422 | 0.02507414397 4117013 |
| **PH PNA_average** | 104179400.0 | 0.01509547699047 7558 | 0.06274660135 590827 | 0.08502696144 513348 |
| **PO2 PNA_average** | 104181846.0 | 0.01590161858610 3467 | 0.08315408369 715689 | 0.09798463197 627395 |
| **LAKTAT_average** | 104832485.0 | 0.01827946671261 8517 | 0.01633610713 8886086 | 0.01511024811 4427134 |
| **Positive_tunge_test** | 104279640.0 | 0.02010326812409 71 | 0.00493380186 70358145 | 0.00700997573 469938 |
| **ICD_Y** | 104306232.0 | 0.02116846569835 4056 | 0.00330103290 3844106 | 0.00525748180 10245345 |
| **urinarytractinfection** | 105275162.5 | 0.02263243152334 8557 | 0.06978312568 771519 | 0.06228093825 829065 |
| **Positive_blod_test** | 104441769.5 | 0.02318930309133 5756 | 0.00017747488 730344657 | 0.00067403612 83364788 |
| **LAKTAT BLODGASS VENØST** | 104607932.5 | 0.02526301495150 3338 | 0.00239591097 85965287 | 0.00070099757 34699381 |

| | | | | |
|---|---|---|---|---|
| **LAKTAT** | 104596786.5 | 0.037297864554986 76 | 0.00192027828 06232918 | 0.00159072526 28740902 |
| **procedure_H** | 104549292.0 | 0.046843265692264 754 | 0.00063890959 42924076 | 0.0 |
| **Positive_biopsi_test** | 104602027.0 | 0.048815044929333 695 | 0.00212969864 76413587 | 0.00080884335 40037746 |
| **procedure_Q_aggregate** | 103377301.5 | 0.050414064747438 97 | 1.07517836226 174 | 1.08425451604 20598 |
| **skinandsofttissueinfection** | 104985557.0 | 0.064324173436553 13 | 0.04074823412 487133 | 0.03612833647 883527 |
| **Positive_eye_test** | 104686549.5 | 0.071116682285421 4 | 0.00716998544 70592414 | 0.00539228902 6691831 |
| **ICD_T** | 104118058.0 | 0.088458085247506 52 | 0.02679870798 282043 | 0.03302777028 848746 |
| **procedure_5_aggregate** | 104390684.5 | 0.095109039704038 44 | 0.00252014339 9708941 | 0.00417902399 56861685 |
| **LAKTAT PNA** | 104446213.5 | 0.100746336845616 1 | 0.00033365278 813047954 | 0.00155028309 5173901 |
| **PO2 PNA** | 104446235.0 | 0.100900762190010 72 | 0.00325133993 5399141 | 0.00636290105 1496361 |
| **PH PNA** | 104446250.5 | 0.101012208736517 49 | 0.00264118127 2849892 | 0.00593151792 9361013 |
| **Positive_hud_test** | 104173359.0 | 0.101535564882528 85 | 0.02498846413 2325278 | 0.02615260177 945538 |
| **Positive_annet_test** | 104842672.0 | 0.116360630448408 72 | 0.03439463315 9407946 | 0.02938797519 5470477 |
| **centralnervoussystem** | 104539664.0 | 0.174655925357613 87 | 0.00070989954 92137863 | 0.00026961445 133459155 |
| **ICD_X** | 104535955.0 | 0.201491958480011 56 | 0.00067440457 17530969 | 0.00026961445 133459155 |
| **procedure_I** | 104599916.5 | 0.248471365946119 63 | 0.00571469137 1170979 | 0.00431383122 1353465 |
| **ICD_T_aggregate** | 103897761.0 | 0.333648089710753 | 0.94842579774 96184 | 0.80304664330 00809 |
| **procedure_K** | 104682388.5 | 0.353116282693785 9 | 0.02757959748 6955595 | 0.02507414397 4117013 |
| **procedure_Z** | 104279583.0 | 0.373057178264475 26 | 0.03393319845 241898 | 0.03626314370 450256 |

| | | | | |
|---|---|---|---|---|
| **lung** | 104395922.0 | 0.4292301896754306 | 0.00876725943279026 | 0.009301698571043408 |
| **ICD_K** | 104793352.5 | 0.4674863224327619 | 0.10975047030845135 | 0.11957400916689134 |
| **ICD_S** | 104305001.0 | 0.474108183567772 07 | 0.043623327299187165 | 0.05230520355891076 |
| **procedure_B** | 104510733.5 | 0.587741174093966 | 0.000567919639371029 | 0.00053922890266918 31 |
| **CRP-HØYSENSITIV_average** | 104278146.5 | 0.5954081187949578 | 1.5298287915461108 | 3.1688857896394973 |
| **BILIRUBIN KONJUGERT** | 104524746.5 | 0.5961748344073067 | 0.03514002768608242 | 0.12995416554327313 |
| **Positive_melk_test** | 104405552.5 | 0.5997749612039122 | 0.018457388279558444 | 0.02291722836344028 |
| **ICD_X_aggregate** | 104566435.0 | 0.6298775519508257 | 0.03095162034572108 | 0.0319493124831491 |
| **procedure_D_aggregate** | 104264702.5 | 0.6753327193466161 | 1.080076669151315 | 1.0532488541385818 |
| **ICD_V** | 104507782.0 | 0.6835816332286178 | 0.0006744045717530969 | 0.0005392289026691831 |
| **endocarditis** | 104478898.0 | 0.7580835652481792 | 0.0012068292336634367 | 0.0013480722566729577 |
| **BILIRUBIN UKONJUGERT** | 104504153.0 | 0.8303720964799104 | 0.09210946651048876 | 0.036397950930169856 |
| **Positive_anus_test** | 104461411.0 | 0.8666174357440665 | 0.022858765484683918 | 0.021838770558101916 |
| **procedure_N** | 104467335.0 | 0.9012484926199882 | 0.035707947325453446 | 0.04044216770018873 |
| **ppid** | 104557209.0 | 0.9356647615510758 | 18170.396052958506 | 18159.356834726343 |
| **Positive_bein_test** | 104494406.5 | 0.9630362622199063 | 0.00014197990984275724 | 0.00013480722566729578 |

**Supplementary Table 5. Sorted statistical summary table for 30-day mortality predictors at the start of the episode.**

| | Stat | P-Value | Mean (Class 0) | Mean (Class 1) | Chi2 Stat | Proportion (Class 0) | Proportion (Class 1) |
|---|---|---|---|---|---|---|---|

| | | | | | | | |
|---|---|---|---|---|---|---|---|
| **ICD_C** | 71549881.0 | 0.0 | 0.10201256522202108 | 0.6658128875707738 | | | |
| **CRP** | 64524392.5 | 0.0 | 30.335534021935885 | 74.67476633414219 | | | |
| **LEUKOCYTTER** | 74426042.0 | 0.0 | 6.576182751807295 | 10.83327446751144 | | | |
| **ICD_I** | 78565754.5 | 0.0 | 0.20370567564689596 | 0.746157994068482 | | | |
| **ICD_J** | 80707904.0 | 0.0 | 0.14293827423419586 | 0.49015907252628743 | | | |
| **procedure_R_aggregate** | 80991734.0 | 0.0 | 1.203457210804671 | 1.8061472094904287 | | | |
| **ICD_J_aggregate** | 75051088.0 | 0.0 | 2.1058460227877753 | 4.130493394445942 | | | |
| **ICD_I_aggregate** | 70409711.5 | 0.0 | 5.215170553366699 | 8.984631976273928 | | | |
| **age** | 55954082.5 | 0.0 | 55.44148652965605 | 75.13345915341063 | | | |
| **total_los** | 55653342.5 | 0.0 | 29.37551319821564 | 55.32891839669267 | | | |
| **time_to_last** | 139810218.5 | 0.0 | 4029.568381074078 | 1528.2231059584794 | | | |
| **care_level_code** | | 0.0 | | | 5309.82522571323 | 0.702090654172434 5 | 0.6952008627662443 |
| **urgency_code** | | 0.0 | | | 3476.8719636480846 | 0.688744542647215 4 | 0.6350768401186303 |
| **ICD_C_aggregate** | 66088361.5 | 0.0 | 5.717992404074823 | 21.923159881369642 | | | |
| **procedure_G** | 92445021.5 | 6.461147808177588e-307 | 0.05373939587548362 | 0.2100296575896468 | | | |
| **ICD_D_aggregate** | 81522390.0 | 1.0225342241721986e-248 | 1.3226138501402052 | 2.241978970072796 | | | |
| **KREATININ_average** | 78101941.5 | 1.7937638791577205e-246 | 69.55719831856099 | 96.03102039289094 | | | |
| **BILIRUBIN TOTAL** | 84048749.5 | 2.224428000205524e-239 | 4.395248405683929 | 10.34822953176957 | | | |
| **KREATININ** | 79927593.0 | 9.871251480293905e-217 | 57.850612051728 | 95.90825536982118 | | | |
| **procedure_G_aggregate** | 84138332.5 | 2.5276146830778575e-213 | 1.2656799062932595 | 2.038285252089512 | | | |
| **ICD_E_aggregate** | 83236161.5 | 2.527435546142632e-204 | 2.262733823164022 | 2.9621191695874898 | | | |
| **procedure_U_aggregate** | 84930257.0 | 9.117993254886273e-203 | 0.7292798069073226 | 1.3982205446211917 | | | |
| **prior_comorbidities_counts** | 82260989.5 | 1.0661305606478129e-197 | 0.88620310226103 | 1.2905095713130224 | | | |

| | | | | |
|---|---|---|---|---|
| **cancer** | 93554126.5 | 8.975823485430375e-180 | 0.06680154758101729 | 0.18172014019951468 |
| **procedure_R** | 98428871.0 | 3.090305100186988e-166 | 0.015582295105242608 | 0.08344567268805608 |
| **Positive_urin_test** | 85159359.0 | 1.7940187801159525e-161 | 1.4234905760834842 | 2.040442167700189 |
| **procedure_P_aggregate** | 90861981.0 | 9.09574276985113e-142 | 0.33454016256699676 | 0.6221353464545699 |
| **cardiovascular** | 89793234.5 | 1.5143070950098356e-134 | 0.24228871614666525 | 0.3968724723645187 |
| **procedure_W_aggregate** | 86694332.0 | 4.942459850275391e-125 | 3.9896354665814786 | 7.32704232946886 |
| **ICD_N** | 93989762.0 | 1.2466367731610224e-119 | 0.11560714158946508 | 0.23726071717444056 |
| **ICD_E** | 95326940.5 | 6.173932779419837e-118 | 0.08564938061264331 | 0.20490698301428956 |
| **procedure_T_aggregate** | 90388872.0 | 3.177158173687538e-113 | 0.6153764242359706 | 1.0806147209490429 |
| **ICD_N_aggregate** | 88194014.5 | 5.312688476904842e-110 | 3.7141589465090687 | 7.733620922081424 |
| **procedure_P** | 101035132.0 | 1.8426844194823795e-108 | 0.007631420154048202 | 0.05217039633324346 |
| **procedure_Y_aggregate** | 102541509.5 | 8.605980998851064e-105 | 0.0004969296844496504 | 0.028174710164464815 |
| **procedure_Y** | 102736544.5 | 2.316684852241728e-104 | 3.549497746068931e-05 | 0.02386087894311135 |
| **ICD_O_aggregate** | 110650265.0 | 1.5626028601862474e-96 | 0.49135697298832215 | 0.009706120248045295 |
| **procedure_M_aggregate** | 110566746.5 | 8.144818502358003e-96 | 0.46778830795442444 | 0.012671879212725803 |
| **procedure_F_aggregate** | 90623111.5 | 7.971645053399038e-94 | 1.4430838036417848 | 1.916149905634942 |
| **Positive_edta_test** | 96825745.5 | 4.626669279080359e-92 | 0.121215348028254 | 0.2636829334052305 |
| **ICD_A_aggregate** | 92787483.5 | 4.850371328140499e-89 | 0.38441060589926523 | 0.5923429495820977 |
| **ICD_Z** | 93957829.0 | 6.706717631485326e-82 | 0.19075000887374435 | 0.3025074143974117 |
| **Positive_blood_culture_test_test** | 96757959.5 | 1.4437130155803911e-74 | 0.11180917900117134 | 0.22661094634672418 |

| | | | | |
|---|---|---|---|---|
| **total_ICU_LOS** | 90616578.5 | 9.549015002490682e-74 | 1.8076231379926433 | 2.9272321829783507 |
| **ICD_A** | 99770196.5 | 1.52009926562003e-71 | 0.03041919568381074 | 0.0764356969533567 |
| **ICD_D** | 99455368.0 | 5.680102566866028e-67 | 0.03911546516167962 | 0.09989215421946616 |
| **PH_average** | 110946234.0 | 2.497893010108104e-65 | 0.711623004791627 | 0.2570426635391474 |
| **procedure_T** | 102468272.0 | 1.458061868973886e-62 | 0.004188407340361339 | 0.025208951199784307 |
| **Positive_hal_test** | 99488660.0 | 3.2377610744579957e-62 | 0.05040286799417883 | 0.11674305742787813 |
| **ICD_P_aggregate** | 109021797.0 | 6.802837309796926e-62 | 0.3350725872289071 | 0.04286869776220006 |
| **organdysfunction** | 97949570.0 | 1.6610888574467235e-57 | 0.09239342633017428 | 0.15772445403073604 |
| **ICD_Z_aggregate** | 92240451.5 | 1.6101120291431334e-55 | 8.39953146629752 | 13.943246157994068 |
| **LAKTAT BLODGASS_average** | 108309454.0 | 2.352992528384936e-52 | 0.08016756706157388 | 0.014720721913844152 |
| **Positive_plasma_test** | 98160723.5 | 1.1178466511556898e-51 | 0.15003726972633372 | 0.26934483688325694 |
| **ICD_M** | 109881934.0 | 1.711999836259085e-51 | 0.09125758705143222 | 0.03531949312483149 |
| **ICD_R_aggregate** | 92940110.0 | 2.996441892051741e-51 | 2.7780854009157703 | 2.8854138581827984 |
| **ICD_H** | 108544372.5 | 1.7543104142969675e-49 | 0.05530117488375395 | 0.011863035858722027 |
| **ICD_F** | 100167796.0 | 6.506232912619054e-47 | 0.048805594008447806 | 0.09355621461310326 |
| **CRP_average** | 93391150.5 | 3.456787092532515e-45 | 54.92968674355863 | 60.5767158248939 |
| **PH** | 107810685.5 | 2.0896554961794823e-44 | 0.27922538127521584 | 0.04523860878943112 |
| **TROMBOCYTTER** | 94768752.5 | 5.035339015328122e-38 | 140.32223878654509 | 163.23831670710885 |
| **procedure_X_aggregate** | 101497771.5 | 3.882770842424184e-37 | 0.02903489156284386 | 0.0637638177406309 |
| **infection** | 96302059.5 | 4.3075193822821885e-35 | 0.396549888190821 | 0.49770827716365595 |

| | | | | | | | |
|---|---|---|---|---|---|---|---|
| **Positive_bronki_test** | 102283452.5 | 7.178507778505122e-34 | 0.015724275015085366 | 0.04017255324885414 | | | |
| **ICD_Q_aggregate** | 108181231.0 | 2.187826908231105e-32 | 0.4477691406665957 | 0.1275276354812618 | | | |
| **dementia** | 102049888.0 | 1.430375398988515e-31 | 0.019912682355446704 | 0.04367754111620383 | | | |
| **BILIRUBIN TOTAL_average** | 95565379.5 | 3.690748862302253e-30 | 13.487312738558972 | 12.612933964125258 | | | |
| **procedure_D** | 106844171.0 | 1.1024833607018874e-28 | 0.04319738756965889 | 0.009301698571043408 | | | |
| **procedure_C** | 106377213.5 | 3.708607221870702e-28 | 0.027047172825045256 | 0.0018873011593421407 | | | |
| **ICD_F_aggregate** | 98508343.5 | 6.300614687658106e-26 | 3.6310297092961346 | 2.371528713939067 | | | |
| **ICD_K_aggregate** | 96903020.5 | 3.4506788715844646e-25 | 2.7748908529443086 | 2.577514154758695 | | | |
| **procedure_U** | 103359168.5 | 1.0006194954820554e-23 | 0.004756326979732368 | 0.016985710434079267 | | | |
| **procedure_O** | 106600839.5 | 1.0802149080806325e-23 | 0.039186455116601 | 0.010245349150714478 | | | |
| **ICD_O** | 105716330.0 | 4.987008878310634e-20 | 0.020232137152592908 | 0.0004044216770018873 | | | |
| **procedure_L_aggregate** | 108312258.5 | 3.421784574536348e-19 | 0.3592446668796365 | 0.20463736856295497 | | | |
| **ICD_R** | 100540188.0 | 5.245791812563757e-19 | 0.12011500372697263 | 0.16486923699110273 | | | |
| **CRP-HØYSENSITIV** | 105832562.5 | 1.288711929787807e-18 | 0.11413161537642413 | 0.187502022108385 | | | |
| **LAKTAT BLODGASS VENØST_average** | 105644995.5 | 6.1576740822351074e-18 | 0.02119061140943795 | 0.0038532398669902036 | | | |
| **Positive_naso_test** | 101077106.5 | 1.700408356522734e-17 | 0.11716892059773543 | 0.16122944189808575 | | | |
| **procedure_M** | 105483960.0 | 3.89391677880088e-17 | 0.016185709722074327 | 0.0 | | | |
| **procedure_W** | 101404125.0 | 2.4533262939378747e-16 | 0.08607532034217158 | 0.1365597196009706 | | | |
| **procedure_Z_aggregate** | 110298205.0 | 3.5800802194477833e-16 | 3.829872573030916 | 2.597600431383122 | | | |
| **Gender** | | 3.3766183934485417e-15 | | | 62.033511631887876 | 0.513967273630 7812 | 0.5653815044486384 |

| | | | | |
|---|---|---|---|---|
| **ICD_L** | 106464572.5 | 1.7570879969753303e-14 | 0.044617186668086464 | 0.023321650040442166 |
| **ICD_B_aggregate** | 100296963.5 | 1.3652839047064898e-13 | 0.5155290526390516 | 0.5246697222971152 |
| **procedure_K_aggregate** | 100408301.0 | 2.1931128269290067e-13 | 1.5470485926241437 | 4.92922620652467 |
| **ICD_H_aggregate** | 100033294.5 | 8.440886746405153e-13 | 1.9927590245980193 | 2.0548665408465894 |
| **TROMBOCYTTER_average** | 110046734.0 | 1.7144484165889526e-12 | 235.96674330079503 | 229.28658489007347 |
| **ICD_W_aggregate** | 102595212.0 | 1.8070273855213364e-12 | 0.041529123629006494 | 0.06457266109463468 |
| **ICD_Y_aggregate** | 102294515.5 | 1.0684703916042478e-11 | 0.07024456039470416 | 0.09112968455109194 |
| **ICD_M_aggregate** | 109235060.5 | 1.5408889040072222e-11 | 3.5540056082064386 | 2.1884605014828793 |
| **sepsis** | 103765887.0 | 1.8332224667866904e-11 | 0.004898306889575125 | 0.011863035858722027 |
| **procedure_L** | 105175953.0 | 5.234422414085725e-11 | 0.010471018350903347 | 0.0012132650310056618 |
| **LAKTAT BLODGASS** | 105309819.5 | 2.4410928420978166e-10 | 0.01990972444065831 | 0.009146670261526015 |
| **procedure_O_aggregate** | 108276514.0 | 2.19729290443262e-09 | 3.365704752777482 | 2.3685629549743865 |
| **procedure_F** | 102957228.5 | 5.169091803468203e-09 | 0.04131615376424236 | 0.07400916689134537 |
| **ICD_G** | 102650295.0 | 6.364643323736832e-09 | 0.05583359954566429 | 0.07764896198436236 |
| **LEUKOCYTTER_average** | 99950989.0 | 7.905770560694441e-09 | 8.751987631339986 | 9.278849674972538 |
| **procedure_I_aggregate** | 106982345.5 | 1.5476062983633245e-08 | 1.6968373975082527 | 0.9770827716365598 |
| **procedure_A** | 103420001.5 | 2.9039856170592678e-08 | 0.028147517126326623 | 0.040711782151523324 |
| **procedure_C_aggregate** | 101684380.5 | 7.145732828447206e-08 | 1.0571469137117098 | 1.059719600970612 |
| **procedure_E** | 104999648.0 | 7.37792037756632e-08 | 0.00724097540198062 | 0.0010784578053383662 |
| **procedure_B_aggregate** | 105417300.5 | 3.5983887950573667e-07 | 0.034785077911475525 | 0.016042059854408198 |
| **procedure_J** | 106061536.5 | 5.209436631655601e-07 | 0.07123841976360346 | 0.06848207063898624 |

| | | | | |
|---|---|---|---|---|
| **ICD_G_aggregate** | 101544903.5 | 8.105848268610126e-07 | 1.64366592127214 | 1.4228902669183068 |
| **explicitsepsis** | 104208325.0 | 1.3854254924359366e-06 | 0.0013843041209668833 | 0.004044216770018873 |
| **ICD_Q** | 105217534.5 | 1.7769583708007634e-06 | 0.01448195080396124 | 0.009301698571043408 |
| **Positive_ear_test** | 105050587.5 | 7.654135066836614e-06 | 0.013665566322365385 | 0.006605554057697492 |
| **procedure_E_aggregate** | 105927272.0 | 9.63951782672537e-06 | 0.11141873424910376 | 0.08708546778107307 |
| **ICD_U** | 104190510.5 | 1.072528293856268e-05 | 0.001952223760337912 | 0.004987867349689944 |
| **ICD_U_aggregate** | 103843306.5 | 2.322706947488587e-05 | 0.02541440386185355 | 0.06484227554596926 |
| **procedure_Q** | 105256300.0 | 3.257405883344266e-05 | 0.023994604763425974 | 0.01712051765974656 |
| **ICD_P** | 105030545.5 | 3.3008008191774997e-05 | 0.01803144855003017 | 0.019681854947425182 |
| **LAKTAT PNA** | 104297841.5 | 9.12506704269565e-05 | 0.0009122209207397154 | 0.004650849285521705 |
| **PH PNA** | 104298059.5 | 9.291138991212599e-05 | 0.007130940971852482 | 0.020942302507414398 |
| **PO2 PNA** | 104298089.0 | 9.31383026386289e-05 | 0.009512653959464734 | 0.02310595847937449 |
| **ICD_S_aggregate** | 101948491.0 | 9.759260890999675e-05 | 1.0059631562133957 | 0.9421677001887301 |
| **procedure_A_aggregate** | 106855752.5 | 0.00010726486312645504 | 0.9360380506158379 | 0.8627662442706929 |
| **LAKTAT BLODGASS VENØST** | 104725861.0 | 0.00024747469371452525 | 0.004467042913427751 | 0.002709625235912645 |
| **ICD_V_aggregate** | 104926858.0 | 0.0006810252641772957 | 0.019948177332907394 | 0.021838770558101916 |
| **procedure_H_aggregate** | 105171323.0 | 0.0007212888704293516 | 0.043126397614737515 | 0.02480452952278242 |
| **ICD_B** | 103712851.5 | 0.0018690745578963171 | 0.035211017641003796 | 0.04543003504987867 |
| **procedure_J_aggregate** | 102456777.5 | 0.0027657524115532192 | 1.3723068185851701 | 1.5163116743057428 |
| **procedure_X** | 104434351.5 | 0.003718840069792487 | 0.00010648493238206793 | 0.0006740361283364788 |
| **procedure_N_aggregate** | 106353592.5 | 0.004506992072759957 | 1.197990984275725 | 0.9684551091938528 |
| **ICD_L_aggregate** | 102845052.5 | 0.005577842351054158 | 1.6548113441947965 | 1.4595578322998113 |

| | | | | |
|---|---|---|---|---|
| **PO2_average** | 104465484.0 | 0.00585046974121691 | 0.0 | 0.002709625235912645 |
| **Positive_tunge_test** | 104249969.0 | 0.006651533235239747 | 0.004614347069889611 | 0.00700997573469938 |
| **intraabdominalinfection** | 104605676.5 | 0.008772158504936182 | 0.0012068292336634367 | 0.00013480722566729578 |
| **LAKTAT_average** | 104859529.0 | 0.009680545460867954 | 0.016022335639444014 | 0.014193558979889524 |
| **ICD_W** | 104328414.5 | 0.013045389208181941 | 0.00205870869271998 | 0.0037746023186842814 |
| **pneumonia** | 104010138.0 | 0.01430067147449932 | 0.020516096972278422 | 0.025074143974117013 |
| **LAKTAT PNA_average** | 104184430.5 | 0.01629405722340584 | 0.011095729954211483 | 0.015175923429495825 |
| **PH PNA_average** | 104189654.0 | 0.018372021599011575 | 0.06248145387427692 | 0.08403882448099219 |
| **PO2 PNA_average** | 104192080.0 | 0.019324253927759633 | 0.08276718844283537 | 0.09737799946077112 |
| **ICD_Y** | 104306232.0 | 0.021168465698354056 | 0.003301032903844106 | 0.0052574818010245345 |
| **urinarytractinfection** | 105275162.5 | 0.022632431523348557 | 0.06978312568771519 | 0.06228093825829065 |
| **Positive_led_test** | 104637665.0 | 0.02762038785865171 | 0.004259397295282717 | 0.0016176867080075492 |
| **Positive_annet_test** | 104948116.5 | 0.03513862697818076 | 0.03247790437653072 | 0.026557023456457267 |
| **procedure_H** | 104549292.0 | 0.046843265692264754 | 0.0006389095942924076 | 0.0 |
| **procedure_Q_aggregate** | 103377301.5 | 0.05041406474743897 | 1.07517836226174 | 1.0842545160420598 |
| **PO2** | 104479570.5 | 0.0513244353821535 | 0.0 | 0.0014289565920733351 |
| **Positive_biopsi_test** | 104594609.0 | 0.06179343151308095 | 0.00205870869271998 | 0.0008088433540037746 |
| **skinandsofttissueinfection** | 104985557.0 | 0.06432417343655313 | 0.04074823412487133 | 0.03612833647883527 |
| **BILIRUBIN UKONJUGERT_average** | 104194926.5 | 0.07129469009298285 | 0.6289149666851721 | 0.4133054731733621 |
| **ICD_T** | 104118058.0 | 0.08845808524750652 | 0.02679870798282043 | 0.03302777028848746 |
| **LAKTAT** | 104573063.5 | 0.13822830903574057 | 0.0021722926205941864 | 0.0024534915071447827 |
| **Positive_eye_test** | 104645015.5 | 0.14148421015123047 | 0.006602065807688212 | 0.0052574818010245345 |
| **BILIRUBIN KONJUGERT_average** | 104239162.0 | 0.1678381341728712 | 0.6198953063051666 | 1.1386739462568525 |
| **Positive_anus_test** | 104744105.5 | 0.17244461147737822 | 0.021438966386256343 | 0.01725532488541386 |
| **centralnervoussystem** | 104539664.0 | 0.17465592535761387 | 0.0007098995492137863 | 0.00026961445133459155 |
| **BILIRUBIN KONJUGERT** | 104575173.5 | 0.17726409108466723 | 0.05327796116849466 | 0.10380156376381774 |
| **Positive_faeces_test** | 104200150.0 | 0.1855660250464709 | 0.03624037198736379 | 0.03585872202750067 |

| | | | | |
|---|---|---|---|---|
| **ICD_X** | 104535955.0 | 0.20149195848001156 | 0.0006744045717530969 | 0.00026961445133459155 |
| **Positive_hud_test** | 104271226.5 | 0.2378165354338586 | 0.023391190146594257 | 0.023726071717444053 |
| **Positive_blod_test** | 104469942.5 | 0.24603295073416176 | 0.00017747488730344657 | 0.0004044216770018873 |
| **procedure_I** | 104599916.5 | 0.24847136594611963 | 0.005714691371170979 | 0.004313831221353465 |
| **BILIRUBIN UKONJUGERT** | 104544197.5 | 0.33188717359774134 | 0.09058318247967913 | 0.02601779455378808 |
| **ICD_T_aggregate** | 103897761.0 | 0.333648089710753 | 0.9484257977496184 | 0.8030466433000809 |
| **procedure_K** | 104682388.5 | 0.3531162826937859 | 0.027579597486955595 | 0.025074143974117013 |
| **procedure_Z** | 104279583.0 | 0.37305717826447526 | 0.03393319845241898 | 0.03626314370450256 |
| **lung** | 104395922.0 | 0.4292301896754306 | 0.00876725943279026 | 0.009301698571043408 |
| **ICD_K** | 104793352.5 | 0.4674863224327619 | 0.10975047030845135 | 0.11957400916689134 |
| **ICD_S** | 104305001.0 | 0.47410818356777207 | 0.043623327299187165 | 0.05230520355891076 |
| **Positive_melk_test** | 104588050.0 | 0.5587532011889409 | 0.017392538955737762 | 0.019681854947425182 |
| **procedure_B** | 104510733.5 | 0.587741174093966 | 0.000567919639371029 | 0.0005392289026691831 |
| **ICD_X_aggregate** | 104566435.0 | 0.6298775519508257 | 0.03095162034572108 | 0.0319493124831491 |
| **procedure_D_aggregate** | 104264702.5 | 0.6753327193466161 | 1.080076669151315 | 1.0532488541385818 |
| **ICD_V** | 104507782.0 | 0.6835816332286178 | 0.0006744045717530969 | 0.0005392289026691831 |
| **CRP-HØYSENSITIV_average** | 104644460.0 | 0.7023105934003187 | 1.4005240259616638 | 2.4888569392883575 |
| **endocarditis** | 104478898.0 | 0.7580835652481792 | 0.0012068292336634367 | 0.0013480722566729577 |
| **procedure_N** | 104467335.0 | 0.9012484926199882 | 0.035707947325453446 | 0.04044216770018873 |
| **ppid** | 104557209.0 | 0.9356647615510758 | 18170.396052958506 | 18159.356834726343 |
| **Positive_bein_test** | 104494406.5 | 0.9630362622199063 | 0.00014197990984275724 | 0.00013480722566729578 |

### Supplementary Table 6. Statistical summary table for readmission predictors.

| | Stat | P-Value | Mean (Class 0) | Mean (Class 1) | Chi2 Stat | Proportion (Class 0) | Proportion (Class 1) |
|---|---|---|---|---|---|---|---|
| **urgency_code** | | 0.0 | | | 1495.6132847379947 | 0.6807283763277694 | 0.49494609164420483 |
| **care_level_code** | | 0.0 | | | 1675.9580309925614 | 0.6700050581689428 | 0.503032345013477 |
| **readmission** | 73300270.0 | 1.5617851875270433e-259 | 0.10706457595683695 | 0.2742587601078167 | | | |

| | | | | |
|---|---|---|---|---|
| **CRP** | 64015954.5 | 1.3346958372170159e-253 | 24.322300904850223 | 52.30337488769092 |
| **LOS** | 65080854.5 | 2.7310169368787146e-250 | 1.5301312313831048 | 2.154537286612745 |
| **age** | 64884344.0 | 5.841356260310446e-225 | 55.242454897993596 | 67.17553908355795 |
| **ICD_C** | 73912389.5 | 2.718255576854275e-215 | 0.15420671050413085 | 0.37331536388140163 |
| **ICD_C_aggregate** | 71184081.5 | 5.6036103992561625e-192 | 7.42400944191536 | 16.129885444743934 |
| **LEUKOCYTTER** | 68683534.0 | 9.318289074767002e-162 | 5.750231607935724 | 8.497265554806827 |
| **total_los** | 70764668.0 | 5.3713693944255296e-126 | 30.803217557466315 | 42.78442273135668 |
| **ICD_I_aggregate** | 71841426.0 | 2.090376144569754e-124 | 5.235238576968471 | 7.926212938005391 |
| **time_to_last** | 104469491.5 | 7.37950298752333e-115 | 2926.7478334176362 | 2177.162398921833 |
| **ICD_J** | 79396887.0 | 1.0660688098700882e-102 | 0.12409374473107401 | 0.26398247978436656 |
| **procedure_0_aggregate** | 82334880.0 | 3.569048563069573e-101 | 0.3897824987354578 | 1.3396226415094339 |
| **ICD_I** | 78510831.0 | 2.7391505957388863e-92 | 0.22094081942336874 | 0.44845013477088946 |
| **procedure_R_aggregate** | 78542812.5 | 2.840466792866408e-88 | 1.2972854493340078 | 1.3096361185983827 |
| **KREATININ_average** | 73650858.0 | 3.732200210925262e-88 | 67.64950820517976 | 83.89065381999715 |
| **ICD_J_aggregate** | 76406340.0 | 9.980969169546653e-69 | 2.094587759231158 | 3.401280323450135 |
| **prior_comorbidities_counts** | 76643264.0 | 5.989805463824227e-64 | 0.8601584892935424 | 1.1159029649595686 |
| **ICD_D_aggregate** | 77890238.5 | 2.601211618606366e-60 | 1.360816051256112 | 1.9701819407008085 |
| **ICD_E_aggregate** | 77796466.5 | 1.2492676441250163e-59 | 2.2152419490810993 | 2.711421832884097 |
| **KREATININ** | 76496210.0 | 2.256786006386439e-59 | 52.82314058899562 | 71.55912314690026 |
| **cancer** | 82571646.0 | 4.603968907920082e-55 | 0.07752486933063564 | 0.14487870619946092 |
| **BILIRUBIN TOTAL** | 80492324.0 | 3.238969065590062e-53 | 4.107008374079694 | 6.576819407008086 |
| **cardiovascular** | 79751986.0 | 4.767010405249556e-53 | 0.24535491485415614 | 0.3507412398921833 |
| **Positive_urin_test** | 78606541.0 | 1.4163923017114096e-47 | 1.393019726858877 | 1.7946428571428572 |
| **procedure_U_aggregate** | 79907768.0 | 4.255295383284669e-43 | 0.8004046535154274 | 1.1661051212938005 |
| **CRP_average** | 78219377.0 | 5.63306620682026e-42 | 48.71917617114914 | 56.517417597312715 |
| **ICD_N_aggregate** | 79394841.5 | 5.5224086480021174e-39 | 4.225324565840499 | 5.3382749326145555 |
| **procedure_P_aggregate** | 82215168.0 | 5.122221292510266e-33 | 0.3509357612544259 | 0.5106132075471698 |
| **procedure_R** | 85772847.5 | 8.033711873933326e-33 | 0.020367560276513236 | 0.049528301886792456 |

| | | | | |
|---|---|---|---|---|
| **procedure_W_aggregate** | 80167327.5 | 8.52787354188764e-31 | 4.295026133872871 | 6.055424528301887 |
| **ICD_N** | 83911042.0 | 6.278183612780519e-27 | 0.10655875906255269 | 0.16088274932614555 |
| **Positive_edta_test** | 84359958.0 | 1.1093842645937956e-26 | 0.1314112291350531 | 0.22287735849056603 |
| **LEUKOCYTTER_average** | 80499175.0 | 2.2045089545030145e-25 | 8.013050858213386 | 8.927622029236018 |
| **procedure_G_aggregate** | 81996046.0 | 7.355192251622003e-25 | 1.2796493002866296 | 1.6438679245283019 |
| **total_ICU_LOS** | 80942884.5 | 3.033439717215107e-24 | 1.8688276288427987 | 2.3831143306379077 |
| **ICD_H** | 90677588.5 | 5.501753046529051e-23 | 0.05823638509526218 | 0.02543800539083558 |
| **ICU_LOS** | 81336199.0 | 3.3942353360533694e-22 | 0.7634055527454642 | 1.075584007187786 |
| **Positive_hal_test** | 85406185.5 | 8.311769372805821e-22 | 0.05489799359298601 | 0.09838274932614555 |
| **procedure_T_aggregate** | 82642037.0 | 3.696210704360028e-21 | 0.6612712864609678 | 0.9161051212938005 |
| **ICD_P_aggregate** | 90209478.5 | 1.7705250796483557e-20 | 0.29549822964087 | 0.057446091644204854 |
| **TROMBOCYTTER** | 82050085.5 | 1.2642240927501483e-19 | 112.9341623110212 | 127.3829655211141 |
| **procedure_F_aggregate** | 82472784.0 | 1.6993092124089638e-19 | 1.4435339740347328 | 1.7434299191374663 |
| **LAKTAT BLODGASS_average** | 90072910.0 | 2.6706788898267067e-19 | 0.07304264705332761 | 0.030073275607481874 |
| **ICD_Z_aggregate** | 81625740.5 | 4.042991288568356e-19 | 9.001180239419996 | 11.038241239892184 |
| **ICD_M** | 90951164.0 | 5.375035891741112e-19 | 0.08679817905918058 | 0.05053908355795148 |
| **PH_average** | 90946531.0 | 2.9884300676044526e-18 | 0.6176578742451743 | 0.3715277739288312 |
| **procedure_G** | 85919758.0 | 3.315186684805863e-17 | 0.057561962569549825 | 0.08473719676549865 |
| **ICD_A_aggregate** | 83717271.0 | 5.675587630959596e-17 | 0.3704940145000843 | 0.4738881401617251 |
| **infection** | 83057230.0 | 7.3560497479346e-17 | 0.3709323891417973 | 0.4467654986522911 |
| **organdysfunction** | 85115494.0 | 1.1848643025836355e-15 | 0.09158657899173832 | 0.12786388140161725 |
| **ICD_R_aggregate** | 82445780.5 | 1.7140210826138334e-15 | 2.631495531950767 | 2.8684299191374665 |
| **ICD_E** | 85382940.5 | 2.5319967542741406e-15 | 0.0830888551677626 | 0.12415768194070081 |
| **procedure_O** | 89664393.5 | 5.163361719300281e-15 | 0.044916540212443096 | 0.015835579514824796 |
| **procedure_C** | 89293672.5 | 2.6944172761936432e-14 | 0.027617602427921092 | 0.007917789757412398 |
| **ICD_Q_aggregate** | 90018782.5 | 4.3344342892915696e-13 | 0.40283257460799193 | 0.19171159029649595 |
| **Positive_blood_culture_test_test** | 85359079.0 | 1.197280061258437e-12 | 0.11718091384252234 | 0.16795822102425875 |

| | | | | | | | |
|---|---|---|---|---|---|---|---|
| **procedure_D** | 89436196.5 | 1.2008087597879019e-12 | 0.04602933737986849 | 0.016340970350404313 | | | |
| **Positive_plasma_test** | 85332056.0 | 1.4753815123662047e-12 | 0.16017534985668522 | 0.21816037735849056 | | | |
| **BILIRUBIN TOTAL_average** | 83087163.0 | 5.276509128431108e-12 | 12.760131229466372 | 11.952399972421341 | | | |
| **ICD_D** | 86276204.0 | 2.2501592832792343e-11 | 0.044545607823301296 | 0.06856469002695417 | | | |
| **ICD_A** | 86716973.0 | 1.1210599359840189e-10 | 0.024919912325071656 | 0.039588948787062 | | | |
| **Gender** | | 1.3232848857261289e-10 | | | 41.27383347983316 | 0.5050075872534142 | 0.5407681940700808 |
| **procedure_J** | 89791491.5 | 2.5104815996225166e-10 | 0.07317484403979092 | 0.04902291105121294 | | | |
| **ICD_O** | 87194609.0 | 2.6056304721421826e-10 | 0.015646602596526723 | 0.02526954177897574 | | | |
| **dementia** | 86931745.0 | 5.480890761370566e-10 | 0.01834429269937616 | 0.03116576819407008 | | | |
| **procedure_P** | 87289003.0 | 7.106615189332912e-10 | 0.009205867475973697 | 0.021731805929919138 | | | |
| **procedure_0** | 87829231.0 | 7.910368308516164e-10 | 0.0002360478839993256 | 0.0023584905660377358 | | | |
| **ICD_F_aggregate** | 84946526.5 | 1.510781523543353e-09 | 3.370257966616085 | 3.0112870619946093 | | | |
| **ICD_B_aggregate** | 85056581.5 | 3.153549686478206e-09 | 0.4675771370763783 | 0.542621293800539 | | | |
| **CRP-HØYSENSITIV** | 88850395.0 | 6.044930220046778e-09 | 0.14437953127634445 | 0.26448618598382745 | | | |
| **procedure_O_aggregate** | 91334139.5 | 8.485669963711247e-09 | 3.240870005058169 | 2.5444743935309972 | | | |
| **ICD_F** | 86612899.0 | 3.042268460629475e-08 | 0.046231664137582194 | 0.06283692722371968 | | | |
| **ICD_K_aggregate** | 84463034.5 | 8.947591688023902e-08 | 2.6250547968302143 | 2.606469002695418 | | | |
| **procedure_A_aggregate** | 90787366.5 | 5.315475851500062e-07 | 0.9134041476985332 | 0.7730795148247979 | | | |
| **procedure_X_aggregate** | 86946140.0 | 6.617840403864482e-07 | 0.033586241780475465 | 0.04834905660377359 | | | |
| **ICD_S** | 86823192.0 | 6.829974207044328e-07 | 0.040161861406170965 | 0.05929919137466307 | | | |
| **procedure_M** | 87459659.0 | 1.3596108131835067e-06 | 0.011970999831394368 | 0.019036388140161724 | | | |
| **procedure_I** | 88488204.0 | 2.62524404434716e-06 | 0.009846568875400438 | 0.0021900269541778976 | | | |
| **procedure_Z** | 89073300.5 | 3.227614742516566e-06 | 0.039217669870173666 | 0.025438005390083558 | | | |
| **ICD_Y_aggregate** | 86714585.0 | 7.011796439142003e-06 | 0.068049232844377 | 0.08507412398921833 | | | |
| **procedure_Z_aggregate** | 90913497.5 | 8.116798941032414e-06 | 3.6532456584049906 | 2.9659703504043127 | | | |
| **Positive_naso_test** | 86439378.5 | 1.2541324555769335e-05 | 0.11832743213623335 | 0.13611859838274934 | | | |
| **Positive_bronki_test** | 87293160.0 | 1.469610037064528e-05 | 0.019119878603945373 | 0.028975741239892182 | | | |
| **procedure_K_aggregate** | 85885606.0 | 2.3938806322046004e-05 | 2.0813690777271963 | 2.942048517520216 | | | |

| | | | | |
|---|---|---|---|---|
| **ICD_W** | 87799676.0 | 0.00010684087121850253 | 0.0015848929354240432 | 0.004043126684636119 |
| **procedure_I_aggregate** | 89452041.5 | 0.00034270837855681984 | 1.6148035744393863 | 1.1746967654986522 |
| **ICD_M_aggregate** | 90298472.5 | 0.000353595907986693 | 3.308919237902546 | 2.594676549865229 |
| **ICD_U_aggregate** | 87541042.0 | 0.00037567892296961735 | 0.029539706626201315 | 0.038578167115902964 |
| **procedure_B_aggregate** | 88593390.5 | 0.00047783332652764237 | 0.03277693474962064 | 0.018025606469002694 |
| **ICD_W_aggregate** | 87190105.5 | 0.0006034058608635023 | 0.04242117686730737 | 0.051381401617250674 |
| **LAKTAT BLODGASS VENØST_average** | 88422123.0 | 0.0006228011346631226 | 0.017864007514993854 | 0.010455694070080864 |
| **procedure_Q** | 88615094.0 | 0.0006629914839629783 | 0.024785027819929185 | 0.018025606469002694 |
| **ICD_Q** | 88449669.5 | 0.0010730238440882854 | 0.012780306862249199 | 0.00774932614555256 |
| **procedure_U** | 87613351.5 | 0.0011114678505941098 | 0.009644242117686731 | 0.014487870619946091 |
| **PH** | 88671386.0 | 0.001127274120110212 | 0.20492120496824565 | 0.15018222147349505 |
| **procedure_A** | 88583457.5 | 0.0012613864901442552 | 0.02977575451020064 | 0.018194070080862535 |
| **procedure_T** | 87651396.0 | 0.0016861443302171 | 0.008430281571404486 | 0.013477088948787063 |
| **procedure_L** | 88312928.5 | 0.0018573187647902528 | 0.00900354071825999 | 0.004380053908355795 |
| **procedure_H_aggregate** | 88580664.0 | 0.0020215291461093566 | 0.04130837969988198 | 0.02628032345013477 |
| **ICD_L** | 88696176.5 | 0.002808841351225713 | 0.039318833249030516 | 0.03099730458221024 |
| **sepsis** | 87743571.5 | 0.0028113573760507183 | 0.0048221210588433655 | 0.007917789757412398 |
| **ICD_S_aggregate** | 86254079.0 | 0.0028365998569720263 | 0.9551846231664137 | 0.9075134770889488 |
| **ICD_T** | 87452418.0 | 0.003808081747193096 | 0.02508851795649975 | 0.032513477088948785 |
| **pneumonia** | 87522314.5 | 0.004248283097941105 | 0.018546619457089866 | 0.024090296495956873 |
| **ICD_H_aggregate** | 86422913.5 | 0.004954625859284079 | 1.953667172483561 | 1.9883760107816713 |
| **LAKTAT_average** | 88347935.0 | 0.009492872293577153 | 0.015885666753068347 | 0.01136137316561845 |
| **LAKTAT BLODGASS** | 88323462.0 | 0.009926726518562285 | 0.01832799415500477 | 0.013569743935309972 |
| **procedure_Y_aggregate** | 88102112.0 | 0.015940804248609813 | 0.0015174506828528073 | 0.0 |
| **procedure_N_aggregate** | 89444905.0 | 0.016097341805435362 | 1.1414938458944528 | 0.9733827493261455 |
| **ICD_G_aggregate** | 86749262.5 | 0.018912948519713653 | 1.5591637160681167 | 1.4278975741239892 |
| **procedure_X** | 87971567.5 | 0.02542722256533641 | 0.0001686056314280897 | 0.0008423180592991914 |

| | | | | |
|---|---|---|---|---|
| **Positive_ear_test** | 88253733.0 | 0.03279185027354361 | 0.012409374473107401 | 0.008254716981132075 |
| **procedure_M_aggregate** | 88577045.0 | 0.03481765569395891 | 0.37356263699207554 | 0.2929582210242588 |
| **ICD_U** | 87897557.5 | 0.03767811542401563 | 0.0018546619457089867 | 0.003200808625336927 |
| **LAKTAT PNA_average** | 87785714.5 | 0.03792048948039781 | 0.010418984994098799 | 0.011455525606469003 |
| **LAKTAT** | 88113985.5 | 0.03855685648993637 | 0.0019996627887371443 | 0.0012466307277628032 |
| **PH PNA_average** | 87787742.0 | 0.03999197743068937 | 0.055837632776934855 | 0.07513055929919137 |
| **PO2 PNA_average** | 87788501.0 | 0.04065774251199606 | 0.07255774742876414 | 0.09417115902964962 |
| **procedure_N** | 87611852.5 | 0.0532730171901935 | 0.03925139099645928 | 0.04969676549865229 |
| **ICD_O_aggregate** | 88531234.0 | 0.054347285224936745 | 0.38034058337548476 | 0.34754043126684636 |
| **Positive_anus_test** | 87701322.0 | 0.05786967646250166 | 0.019355926487944698 | 0.02324797843665768 |
| **explicitsepsis** | 87924205.0 | 0.07127588213792732 | 0.0015174506828528073 | 0.0026954177897574125 |
| **ICD_B** | 87650774.0 | 0.0745022238878583 | 0.028730399595346484 | 0.032681940700808626 |
| **ICD_L_aggregate** | 87064425.5 | 0.07588444291364484 | 1.6059011970999832 | 1.4142520215633423 |
| **procedure_L_aggregate** | 88697555.0 | 0.07819133792763425 | 0.3219693137750801 | 0.301044474393531 |
| **procedure_B** | 88060560.0 | 0.08307750814225415 | 0.0005058168942842691 | 0.0 |
| **procedure_E** | 88146770.0 | 0.08808939207675169 | 0.004687236553700893 | 0.003200808625336927 |
| **ICD_P** | 87817501.0 | 0.09436418791229256 | 0.013960546282245826 | 0.017014824797843667 |
| **procedure_E_aggregate** | 88474772.0 | 0.11289011634082372 | 0.10237733940313606 | 0.09046495956873316 |
| **procedure_Y** | 88051656.0 | 0.12112615677947058 | 0.0008430281571404484 | 0.0 |
| **ICD_V_aggregate** | 88190204.0 | 0.12354072362021452 | 0.01834429269937616 | 0.02644878706199461 |
| **ppid** | 89122658.0 | 0.12564669459657068 | 18206.21025122239 | 17977.680592991914 |
| **ICD_X** | 87965671.5 | 0.12875589714260807 | 0.0006069802731411229 | 0.0011792452830188679 |
| **BILIRUBIN KONJUGERT** | 88093271.0 | 0.13892995109233436 | 0.048187489462148035 | 0.02341644204851752 |
| **LAKTAT BLODGASS VENØST** | 87947882.0 | 0.16561759261579934 | 0.0028595515090204023 | 0.004750673854447439 |
| **BILIRUBIN UKONJUGERT** | 88072503.0 | 0.1940392830587837 | 0.08582026639689766 | 0.017857142857142856 |
| **procedure_F** | 87711387.5 | 0.19529044717492594 | 0.04265722475130669 | 0.054750673854447436 |
| **procedure_C_aggregate** | 87405751.0 | 0.19886734002739692 | 1.0435002529084472 | 0.9978099730458221 |

| | | | | |
|---|---|---|---|---|
| **procedure_K** | 88257767.5 | 0.20563105976211438 | 0.03146181082448154 | 0.028638814016172506 |
| **PO2** | 88004180.0 | 0.20618354778850345 | 0.0003203506997133704 | 0.0017857142857142857 |
| **PO2_average** | 88004180.0 | 0.20618354778850345 | 0.0003203506997133704 | 0.0017857142857142857 |
| **procedure_J_aggregate** | 87288552.5 | 0.23607070692597942 | 1.3163378856853818 | 1.4081873315363882 |
| **CRP-HØYSENSITIV_average** | 87617765.5 | 0.2654821721488153 | 1.4640893065367517 | 2.224602731023469 |
| **ICD_Y** | 87933223.0 | 0.26898329724714587 | 0.0034732760074186477 | 0.004548517520215634 |
| **ICD_G** | 88317845.0 | 0.2827071964360207 | 0.05648288652841005 | 0.05272911051212938 |
| **Positive_eye_test** | 88114345.5 | 0.2872874511680036 | 0.006305850615410555 | 0.00488544474393531 |
| **Positive_tunge_test** | 87942213.0 | 0.3510812241702028 | 0.004552352048558422 | 0.005390835579514825 |
| **ICD_X_aggregate** | 87891930.5 | 0.3618355912235993 | 0.027887371438206035 | 0.04396900269541779 |
| **centralnervoussystem** | 88042777.0 | 0.3790037807614036 | 0.0006407013994267408 | 0.00033692722371967657 |
| **ICD_T_aggregate** | 88505137.0 | 0.37976528571412604 | 0.9032878098128477 | 0.8237870619946092 |
| **Positive_melk_test** | 87892512.0 | 0.4007442109496363 | 0.017096611026808296 | 0.019036388140161724 |
| **skinandsofttissueinfection** | 88207317.0 | 0.412112542310318 | 0.036519979767324226 | 0.034703504043126686 |
| **procedure_H** | 88039803.5 | 0.4230208254178274 | 0.0007418647782835946 | 0.0005053908355795149 |
| **ICD_V** | 88033860.5 | 0.4380057824949589 | 0.0003709323891417973 | 0.00016846361185983828 |
| **ICD_K** | 88302554.5 | 0.44190018413652754 | 0.1080762097454055 | 0.10545822102425877 |
| **Positive_biopsi_test** | 88051764.5 | 0.4633351502519639 | 0.0018209408194233688 | 0.0011792452830188679 |
| **BILIRUBIN KONJUGERT_average** | 88124808.5 | 0.5144208940802919 | 0.6930146201238765 | 0.7162511230907457 |
| **urinarytractinfection** | 88195591.0 | 0.5457448997207798 | 0.06069802731411229 | 0.05862533692722372 |
| **procedure_D_aggregate** | 88301488.0 | 0.5632163501855436 | 1.0305850615410554 | 1.077156334231806 |
| **ICD_R** | 87807835.0 | 0.5946593273371611 | 0.11849603776766145 | 0.12432614555256065 |
| **procedure_W** | 87853178.0 | 0.6107703872755473 | 0.07941325240263025 | 0.08237870619946092 |
| **TROMBOCYTTER_average** | 87659558.5 | 0.6211694732647592 | 218.80814494768194 | 222.34584165442283 |
| **BILIRUBIN UKONJUGERT_average** | 88084285.5 | 0.647637290804868 | 0.6280325304250444 | 0.34199517070979335 |
| **Positive_faeces_test** | 88101985.5 | 0.6676761620977966 | 0.03550834597875569 | 0.0330188679245283 |

| | | | | |
|---|---|---|---|---|
| **intraabdominalinfection** | 88030930.0 | 0.6767134862252595 | 0.0008430281571404484 | 0.0006738544474393531 |
| **lung** | 87969503.5 | 0.6791578297041623 | 0.008632608329118193 | 0.00876010781671159 |
| **ICD_Z** | 88168371.5 | 0.7609910384420818 | 0.21011633788568537 | 0.2120956873315364 |
| **Positive_hud_test** | 88055232.5 | 0.8180324214567738 | 0.023166413758219526 | 0.019878706199460916 |
| **Positive_bein_test** | 88013084.5 | 0.8420913587293486 | 0.00013488450514247176 | 0.00016846361185983828 |
| **Positive_blod_test** | 88019020.5 | 0.8651673486032568 | 0.00020232675771370763 | 0.00016846361185983828 |
| **Positive_led_test** | 88025145.5 | 0.8739460319538371 | 0.0036081605125611194 | 0.002526954177897574 |
| **LAKTAT PNA** | 88013096.0 | 0.893559098563987 | 0.00032709492497049397 | 0.0003537735849056604 |
| **PO2 PNA** | 88013099.0 | 0.8936669377504577 | 0.0027044343281065588 | 0.0026785714285714286 |
| **PH PNA** | 88013102.0 | 0.8937747788343426 | 0.0022559433485078405 | 0.002486522911051213 |
| **procedure_Q_aggregate** | 87957270.5 | 0.9093148056456916 | 1.0701062215477997 | 0.9765835579514824 |
| **Positive_annet_test** | 87997693.5 | 0.9255723338991142 | 0.030382734783341765 | 0.03133423180592992 |
| **endocarditis** | 88019095.5 | 0.9439790784907232 | 0.0012139605462822458 | 0.0011792452830188679 |

**Supplementary Table 7. Statistical summary table for PLOS predictors.**

| | Stat | P-Value | Mean (Class 0) | Mean (Class 1) | Chi2 Stat | Proportion (Class 0) | Proportion (Class 1) |
|---|---|---|---|---|---|---|---|
| **ICD_N** | 95197989.5 | 0.0 | 0.08213337322811086 | 0.31861305624576464 | | | |
| **urgency_code** | | 0.0 | | | 15060.36740925732 | 0.795190185884729 | 0.8995934041111362 |
| **care_level_code** | | 0.0 | | | 20916.57221538238 | 0.8121704005685005 | 1.0 |
| **CRP** | 79372162.0 | 0.0 | 28.70623418234407 | 72.40364053911603 | | | |
| **age** | 83386041.5 | 0.0 | 56.747952275872386 | 67.99446577817936 | | | |
| **ICD_I** | 84367690.0 | 0.0 | 0.16811908591091 | 0.7656426473910097 | | | |
| **ICD_J** | 78685693.5 | 0.0 | 0.09671990126042562 | 0.5734131466004065 | | | |
| **LEUKOCYTTER** | 84763030.0 | 0.0 | 6.650528107117489 | 9.91832580377982 | | | |
| **ICD_E** | 98548802.0 | 0.0 | 0.056214234955305385 | 0.2744522249830585 | | | |
| **ICD_F** | 106542708.0 | 1.2745780204429456e-298 | 0.03018289262071287 | 0.14253444770725096 | | | |
| **BILIRUBIN TOTAL** | 94728350.0 | 4.574200391106898e-282 | 4.801454912667839 | 8.15608764400271 | | | |
| **ICD_B** | 108816160.5 | 1.0223196160422629e-279 | 0.01540935781875304 | 0.10356900835780439 | | | |

| | | | | |
|---|---|---|---|---|
| **ICD_A** | 108370438.5 | 6.312794597103897e-277 | 0.018326663425215994 | 0.10548904449966116 |
| **procedure_G** | 106514862.0 | 2.1319771605649548e-262 | 0.05281071174776527 | 0.18748588208719222 |
| **procedure_R** | 110410605.5 | 2.3871882748127784e-251 | 0.010883793993342558 | 0.08662751298847979 |
| **KREATININ** | 96841681.0 | 1.5650812595997213e-147 | 60.72928613282467 | 81.04289680746933 |
| **ICD_D** | 110840825.0 | 5.36701768876634e-130 | 0.033137599581104835 | 0.10808674045629094 |
| **ICD_Z_aggregate** | 137035771.0 | 7.7865531223005605e-112 | 10.345588510304074 | 7.167494917551389 |
| **procedure_W** | 109376037.5 | 3.2235409976692005e-111 | 0.07020234132475596 | 0.176304495143438 |
| **procedure_Z_aggregate** | 134461366.5 | 4.036035047790224e-100 | 3.988368178928077 | 2.318838942850689 |
| **procedure_A_aggregate** | 132003182.5 | 4.873650626855162e-98 | 1.042824550248719 | 0.5521798057375198 |
| **procedure_P** | 114908585.0 | 7.57862268887255e-96 | 0.007106257246512324 | 0.04653264061441156 |
| **total_los** | 101058314.0 | 9.069419030854631e-95 | 33.26198401715454 | 39.383395452149834 |
| **ICD_C** | 107698966.5 | 3.584919956974932e-90 | 0.15398137412574336 | 0.4174384459001581 |
| **ICD_J_aggregate** | 102611177.5 | 4.893719415120409e-89 | 2.303100572240715 | 3.206460356900836 |
| **procedure_O_aggregate** | 131538439.0 | 2.5163266207155048e-85 | 3.5620675468451957 | 1.937316467133499 |
| **procedure_M** | 115949153.0 | 6.760917548680435e-83 | 0.0025432920671728315 | 0.04382200135531963 |
| **time_to_last** | 133972477.0 | 1.9140843731189273e-77 | 3380.3748737704304 | 3894.319629545968 |
| **ICD_I_aggregate** | 103916774.0 | 2.8905103160342645e-73 | 5.748513296181322 | 6.7627061215269935 |
| **TROMBOCYTTER** | 103857676.0 | 5.629168095094465e-73 | 137.7152516487764 | 167.3941250658836 |
| **ICD_H** | 123538911.0 | 1.7828958375315663e-70 | 0.058084302651755994 | 0.010503727128981252 |
| **ICD_G** | 112654218.0 | 4.3461133769176205e-64 | 0.04720050865841344 | 0.10018070928393946 |
| **procedure_T** | 116259507.0 | 9.3879165765742e-60 | 0.004039346224333321 | 0.022249830585046308 |
| **ICD_R** | 110724681.0 | 7.875897056614064e-59 | 0.1079777087930583 | 0.19426248023492207 |
| **procedure_J_aggregate** | 129921190.0 | 2.7044977772804e-57 | 1.5247783969779707 | 1.0325276711091032 |
| **TROMBOCYTTER_average** | 131369127.0 | 2.3313029955708225e-54 | 240.50388432112962 | 216.66891694531674 |
| **procedure_D** | 121825118.0 | 2.6018103619645317e-53 | 0.04690129782698134 | 0.0036141856787892477 |
| **ICD_M_aggregate** | 129578997.5 | 8.92186923475864e-51 | 3.6551595167745075 | 2.1044725547775016 |
| **PH_average** | 124151535.5 | 6.56448559736074e-47 | 0.7062891269358192 | 0.34687655356278463 |
| **KREATININ_average** | 106348512.0 | 1.1675794572703303e-46 | 73.41531478357591 | 80.08671630165021 |

| | | | | |
|---|---|---|---|---|
| **ICD_P** | 116443247.5 | 2.7155530047661525e-44 | 0.006694842353293189 | 0.0536480686695279 |
| **procedure_O** | 121430659.5 | 7.982073452118447e-43 | 0.041889516400493695 | 0.00677659814772984 |
| **procedure_C** | 120816959.5 | 2.7163297708891187e-41 | 0.02883644387926843 | 0.00056471651231082 |
| **ICD_Y** | 117200816.5 | 3.1323880983157707e-41 | 0.0011968433257283913 | 0.0112943302462164 |
| **ICD_U** | 117379500.5 | 3.440966299590122e-41 | 0.000486217601077159 | 0.008922520894510955 |
| **procedure_D_aggregate** | 126090474.0 | 3.0509426289488542e-40 | 1.1728690578598946 | 0.7773887508470748 |
| **ICD_O** | 116481448.0 | 3.810676065797105e-40 | 0.006021617982570969 | 0.04653264061441156 |
| **procedure_R_aggregate** | 110939991.0 | 3.26003846937548e-39 | 1.4999812993230355 | 0.8129658911226564 |
| **procedure_W_aggregate** | 128579306.5 | 1.200883881827528e-37 | 4.898343120020945 | 4.041676078608538 |
| **ICD_T_aggregate** | 126757144.0 | 1.7854166439199112e-37 | 1.0145865280323147 | 0.6268353286650101 |
| **LAKTAT BLODGASS_average** | 121703031.5 | 6.065096181084578e-36 | 0.07741839739704597 | 0.033637168599254495 |
| **procedure_N** | 115593573.0 | 1.2447722330614733e-34 | 0.024759696301006096 | 0.07273548678563362 |
| **ICD_E_aggregate** | 109338121.0 | 4.539791051429353e-34 | 2.45891461270898 | 2.25626835328665 |
| **ICD_Q_aggregate** | 122378244.0 | 8.602968063673555e-34 | 0.4701350188876837 | 0.11192681274000452 |
| **ICD_K_aggregate** | 127666775.0 | 7.456217274288996e-33 | 2.972285596738602 | 2.0134402529929973 |
| **dementia** | 115751727.5 | 7.546192323698887e-32 | 0.01914949321165426 | 0.04212785181838717 |
| **procedure_Y** | 117356660.5 | 1.3695756937912265e-31 | 0.0022814825896697462 | 0.013214366388073188 |
| **procedure_N_aggregate** | 126390922.0 | 1.0920549880088293e-30 | 1.24460485469574 | 0.8649198102552519 |
| **ICD_L_aggregate** | 125651176.5 | 1.177245308964648e-30 | 1.80689680966451 | 1.0319629545967923 |
| **CRP-HØYSENSITIV** | 120078830.0 | 3.192319758235104e-26 | 0.1292605752328233 | 0.129916421956178 |
| **ICD_T** | 115952214.5 | 8.57550140118662e-25 | 0.022253805587762278 | 0.04574203749717642 |
| **procedure_Q_aggregate** | 124602148.0 | 9.542376654562553e-25 | 1.1845756816396753 | 0.7524282810029366 |
| **ICD_F_aggregate** | 112207274.0 | 2.4853319601980505e-24 | 3.6777125331937017 | 2.434831714479331 |
| **procedure_E_aggregate** | 121797146.0 | 2.4050811800215e-23 | 0.11972173392676815 | 0.06595888863790378 |
| **procedure_L_aggregate** | 122793961.0 | 1.694545352171947e-22 | 0.3632419493585668 | 0.21764174384459 |
| **cardiovascular** | 112345447.0 | 2.155818110915417e-21 | 0.26050043011557017 | 0.31680596340637 |
| **procedure_I_aggregate** | 122737783.0 | 9.77311705373912e-21 | 1.7357594344915286 | 0.9762819064829455 |

| | | | | |
|---|---|---|---|---|
| **ICD_W** | 117708841.5 | 2.0831699120914233e-20 | 0.0010098365560833303 | 0.006663654845267676 |
| **procedure_Y_aggregate** | 117485122.5 | 3.4650890173771513e-20 | 0.003964543516475297 | 0.013214366388073188 |
| **ICD_K** | 114390313.0 | 1.3876663631424787e-19 | 0.0996746082208176 | 0.14840749943528347 |
| **ICD_R_aggregate** | 125740588.5 | 1.6678699294112032e-19 | 2.927254366608071 | 2.4175513892026204 |
| **ICD_S** | 115868271.5 | 5.593435362540414e-19 | 0.03631671466507087 | 0.07296137339055794 |
| **total_ICU_LOS** | 125467493.5 | 2.4080320622161896e-18 | 2.05670512772562 | 1.993477524282812 |
| **ICD_N_aggregate** | 112014922.0 | 3.449504130781622e-16 | 4.9294984478438115 | 3.4116783374745876 |
| **ICD_B_aggregate** | 113779449.0 | 3.047344106840584e-14 | 0.5234319482365262 | 0.49932234018522703 |
| **procedure_K_aggregate** | 122825414.0 | 5.2103648081532813e-14 | 2.465833863185847 | 1.606166704314434 |
| **ICD_P_aggregate** | 120468592.0 | 4.2477551708534963e-13 | 0.3342184987096533 | 0.0928393946238988 |
| **procedure_H_aggregate** | 119889807.5 | 8.673615302860865e-13 | 0.046003665332685045 | 0.019087418116105714 |
| **ICD_C_aggregate** | 113625989.0 | 1.0661444554588713e-12 | 8.72132251187493 | 10.225547775016942 |
| **Positive_urin_test** | 113018471.0 | 1.9917977627281193e-12 | 1.492875042076523 | 1.7308561102326632 |
| **procedure_B_aggregate** | 119677854.0 | 1.0605687257404573e-11 | 0.036690728204361 | 0.013327309690535352 |
| **Positive_blood_culture_test_test** | 115543049.5 | 3.8167042278419564e-10 | 0.1278378277293638 | 0.15958888637903773 |
| **LAKTAT BLODGASS VENØST_average** | 119246134.5 | 5.386184133886086e-10 | 0.02010272014704075 | 0.009950304946916651 |
| **ICD_H_aggregate** | 122295141.0 | 3.1611893061484266e-09 | 2.13744997568912 | 1.6078608538513666 |
| **LAKTAT_average** | 119223664.0 | 1.152058245330432e-08 | 0.01810639331407534 | 0.008196797429912406 |
| **BILIRUBIN TOTAL_average** | 123115302.0 | 1.1654498207264866e-08 | 14.071635401016888 | 10.99022929888353 |
| **infection** | 122341118.0 | 1.727267496832374e-08 | 0.42529079552679805 | 0.3945109555003388 |
| **urinarytractinfection** | 120328517.0 | 7.358697488484514e-08 | 0.07240902120656768 | 0.05556810481138468 |
| **procedure_F** | 116860696.5 | 7.750297588524213e-08 | 0.03968283651868198 | 0.07363903320533093 |
| **LAKTAT PNA** | 118078812.0 | 7.988092900518435e-08 | 0.0009013726296891948 | 0.004077253218884121 |
| **PH PNA** | 118078884.0 | 8.048159682572808e-08 | 0.005544376706436772 | 0.023493336345154728 |
| **PO2 PNA** | 118078941.5 | 8.096443857468697e-08 | 0.006893069529116953 | 0.028811836458098032 |
| **procedure_I** | 118890379.0 | 8.12172023187473e-08 | 0.006283427460074055 | 0.0028235825615541 |
| **Positive_edta_test** | 116227132.0 | 9.80982098562563e-08 | 0.1379361932901971 | 0.190083578043822 |

| | | | | |
|---|---|---|---|---|
| **Positive_annet_test** | 119581045.5 | 1.1708710888154185e-07 | 0.03418483749111718 | 0.022362773887508472 |
| **ICD_M** | 120354319.5 | 1.6275054305295746e-07 | 0.08415304634027752 | 0.06584594533544161 |
| **ICD_W_aggregate** | 116908072.0 | 3.745526202304411e-07 | 0.04304895837229308 | 0.05624576462615767 |
| **Positive_faeces_test** | 119547093.5 | 5.421373292817618e-07 | 0.03979504058046901 | 0.025186356449062572 |
| **procedure_C_aggregate** | 121124123.0 | 6.622121129208306e-07 | 1.1337846430040768 | 0.827874407047662 |
| **skinandsofttissueinfection** | 119761509.0 | 7.986223224331905e-07 | 0.04271234618693197 | 0.030946464874632935 |
| **Positive_ear_test** | 118994121.0 | 2.0189271172738994e-06 | 0.014100310431237611 | 0.006437768240343348 |
| **procedure_U** | 117798451.5 | 2.4876187008266273e-06 | 0.005909413920783932 | 0.011520216851140727 |
| **ICD_A_aggregate** | 115503784.5 | 4.34823895782659e-06 | 0.42487938063357894 | 0.43641292071380167 |
| **ICD_U_aggregate** | 117616050.5 | 4.725322764524521e-06 | 0.03250177656431163 | 0.03704540320758979 |
| **CRP-HØYSENSITIV_average** | 120260252.5 | 6.363461058604621e-06 | 1.5821364758516838 | 1.763917009737119 |
| **ICD_O_aggregate** | 119742479.0 | 1.1661248728231539e-05 | 0.41695029360062835 | 0.31251411791280775 |
| **procedure_M_aggregate** | 119725601.0 | 1.225471584302843e-05 | 0.3984366234057673 | 0.29591145245086964 |
| **cancer** | 116692968.5 | 4.043055995151331e-05 | 0.08688334517709541 | 0.10243957533318274 |
| **ICD_L** | 119440186.0 | 8.456746114730424e-05 | 0.04256274077121592 | 0.03297944431895189 |
| **BILIRUBIN KONJUGERT_average** | 119111476.5 | 0.00014324691828042122 | 0.7934371250796055 | 0.5304794877586836 |
| **ICU_LOS** | 121311716.0 | 0.0002560669292225152 | 0.8228360199972596 | 1.0409372411716018 |
| **procedure_E** | 118725365.0 | 0.0003130360907041235 | 0.006582638291506153 | 0.0040659588886379034 |
| **ICD_X_aggregate** | 118935719.0 | 0.00038147284057993513 | 0.037139544451509146 | 0.013101423085611023 |
| **LEUKOCYTTER_average** | 115413829.5 | 0.00042838870124627717 | 8.811861295371932 | 9.012595321021971 |
| **ICD_Z** | 116307536.0 | 0.000436946418272741 | 0.20589445337921233 | 0.23864919810255253 |
| **ICD_V** | 118235178.5 | 0.00044533591380206683 | 0.0003740135392901223 | 0.0014682629320081318 |
| **Positive_plasma_test** | 119862843.0 | 0.0007745967182708298 | 0.17997531510640685 | 0.15958888637903773 |
| **BILIRUBIN UKONJUGERT_average** | 118948431.5 | 0.0009284026123674081 | 0.6738136174497771 | 0.3126910624199985 |
| **procedure_X** | 118293342.0 | 0.0010391463433691876 | 7.480270785802445e-05 | 0.0006776598147729839 |
| **ICD_Q** | 118874764.5 | 0.0015625111298600254 | 0.014212514493024648 | 0.010955500338829907 |
| **procedure_A** | 118989492.5 | 0.0024212931594477823 | 0.033100198227175824 | 0.02371809351705444 |

| | | | | | | | |
|---|---|---|---|---|---|---|---|
| **procedure_L** | 118698160.0 | 0.002576012972590941 | 0.00957474660582713 | 0.0054212785181838715 | | | |
| **Positive_melk_test** | 118878761.5 | 0.0027728610669722 | 0.019186894565583274 | 0.01389202620284617 | | | |
| **ICD_G_aggregate** | 120216650.5 | 0.003604564275183135 | 1.7143658600441336 | 1.245199909645358 | | | |
| **Positive_eye_test** | 118680417.5 | 0.003956944491768414 | 0.0070314545386542995 | 0.004178902191100068 | | | |
| **procedure_Z** | 117667248.5 | 0.006396134575241677 | 0.03235217114859558 | 0.040659588886379036 | | | |
| **LAKTAT** | 118519199.0 | 0.006727068678707766 | 0.002633055316602461 | 0.0010164897221594759 | | | |
| **CRP_average** | 116183368.0 | 0.009211567797416978 | 54.46089844400552 | 61.076474013775716 | | | |
| **procedure_G_aggregate** | 116577558.5 | 0.010126399797655014 | 1.422111680442832 | 1.4405918229049017 | | | |
| **Gender** | | 0.010630521431399689 | | | 6.526059351565154 | 0.5285933350787299 | 0.5128755364806867 |
| **procedure_B** | 118448812.0 | 0.012107976666971911 | 0.0007480270785802446 | 0.0 | | | |
| **ICD_V_aggregate** | 118704324.5 | 0.012336056690957527 | 0.020907356846317837 | 0.01863564490625706 | | | |
| **Positive_naso_test** | 119341358.0 | 0.0222308511537434 | 0.131465759060478 | 0.11091032301784504 | | | |
| **centralnervoussystem** | 118444297.5 | 0.027342277562341536 | 0.0007854284325092568 | 0.00011294330246216399 | | | |
| **ICD_S_aggregate** | 119886578.0 | 0.02858702554316921 | 1.0404682649511912 | 0.8483171447933138 | | | |
| **ICD_D_aggregate** | 116833410.0 | 0.034885409721731576 | 1.5434416725885478 | 1.4260221368872825 | | | |
| **LAKTAT BLODGASS VENØST** | 118224968.5 | 0.03825551761110994 | 0.00278079066462206 | 0.008086740456290941 | | | |
| **Positive_led_test** | 118496392.5 | 0.05841374183044616 | 0.0042637543479073945 | 0.0020329794443189517 | | | |
| **procedure_J** | 117762025.0 | 0.06991308124321848 | 0.06466694094326214 | 0.0887734357352609 | | | |
| **organdysfunction** | 117591391.0 | 0.07595536612403558 | 0.10401316527658301 | 0.11203975604246669 | | | |
| **PO2** | 118351330.5 | 0.08226683864783566 | 0.0 | 0.0011971990060989383 | | | |
| **Positive_hud_test** | 118694484.5 | 0.10008115737189566 | 0.02498410442458017 | 0.018861531511181386 | | | |
| **procedure_Q** | 118667309.0 | 0.12139735482286573 | 0.02180498934061413 | 0.024847526541676077 | | | |
| **Positive_biopsi_test** | 118452452.0 | 0.12714060868128027 | 0.0019822717582376483 | 0.0012423763270 83804 | | | |
| **BILIRUBIN KONJUGERT** | 118460947.0 | 0.13444379474569762 | 0.060627594718928826 | 0.0734131466004066 | | | |
| **procedure_T_aggregate** | 119355658.0 | 0.1354958655131674 | 0.7169465534652354 | 0.6984413824260222 | | | |
| **PH** | 118730968.0 | 0.14696812999364892 | 0.2360427497475404 | 0.21358952639108517 | | | |
| **Positive_anus_test** | 118641341.5 | 0.15676602305759826 | 0.02128137038560796 | 0.01840975830133273 | | | |

| | | | | |
|---|---|---|---|---|
| **Positive_bein_test** | 118386834.0 | 0.1981619747409854 | 0.00018700676964506115 | 0.0 |
| **LAKTAT PNA_average** | 118196777.5 | 0.22031269114582175 | 0.01162995100422635 | 0.012900948723740686 |
| **PH PNA_average** | 118201781.5 | 0.2350956535605564 | 0.06444515091446323 | 0.0746126044725548 |
| **PO2 PNA_average** | 118202753.5 | 0.23790053673872802 | 0.08373602124396907 | 0.09208267449740233 |
| **procedure_H** | 118395508.5 | 0.30102401130919487 | 0.0005610203089351835 | 0.00033882990738649197 |
| **Positive_hal_test** | 118049786.0 | 0.3250781646483383 | 0.06264726783109549 | 0.0690083578043822 |
| **explicitsepsis** | 118305384.5 | 0.34577524043785324 | 0.0017952649858592587 | 0.0023718093517054437 |
| **PO2_average** | 118355757.0 | 0.41114845963622504 | 0.0003553128623256162 | 0.0011971990060989383 |
| **Positive_blod_test** | 118382319.5 | 0.4180232440332292 | 0.00026180947750308563 | 0.00011294330246216399 |
| **endocarditis** | 118399327.5 | 0.49714012846814815 | 0.0013090473875154 28 | 0.0010164897221594759 |
| **intraabdominalinfection** | 118395075.5 | 0.5042989701858427 | 0.0010472379100123425 | 0.000790603117235148 |
| **BILIRUBIN UKONJUGERT** | 118399246.0 | 0.5331624599533715 | 0.09058607921606762 | 0.03648068669527897 |
| **prior_comorbidities_counts** | 117890973.5 | 0.548139065282932 | 0.9644313124135093 | 0.9887056697537836 |
| **lung** | 118443351.5 | 0.5500220685373689 | 0.00920073306653701 | 0.00790603117235148 |
| **ICD_Y_aggregate** | 118178540.5 | 0.5887821160279978 | 0.07513932004338557 | 0.07296137339055794 |
| **procedure_U_aggregate** | 117996337.5 | 0.5909049612977192 | 0.8632606500355313 | 0.8851366613959792 |
| **sepsis** | 118315266.5 | 0.6680829726032309 | 0.0062460261061450 42 | 0.006663654845267676 |
| **ICD_X** | 118350893.0 | 0.69526997147073 | 0.0005610203089351835 | 0.0006776598147729839 |
| **procedure_P_aggregate** | 118579437.0 | 0.7075120564045225 | 0.39495829749036915 | 0.3930426925683307 |
| **LAKTAT BLODGASS** | 118411030.0 | 0.7356228835802239 | 0.015598234656094553 | 0.023911979519614483 |
| **ppid** | 118105057.0 | 0.7566690917349355 | 18158.179750906984 | 18198.03761011972 |
| **pneumonia** | 118428296.0 | 0.7620970185174154 | 0.021617982570969068 | 0.021007454257962504 |
| **Positive_tunge_test** | 118338929.5 | 0.7874408175199241 | 0.0050117814264876385 | 0.0054212785181838715 |
| **Positive_bronki_test** | 118416143.5 | 0.7907694307663935 | 0.020757751430601788 | 0.021007454257962504 |
| **procedure_F_aggregate** | 118426500.5 | 0.9314486029541019 | 1.6005535400381494 | 1.3639033205330924 |
| **procedure_X_aggregate** | 118380923.5 | 0.948342968715609 | 0.03635411601899989 | 0.03602891348543032 |
| **procedure_K** | 118377710.5 | 0.9520387674417476 | 0.02584433556494745 | 0.030720578269708608 |

### Supplementary Table 8. Disease Groups

| S/No. | Disease | ICD-10 codes |
|---|---|---|
| 1 | **Explicit Sepsis** | 'A021','A207','A217','A227','A241','A267','A282','A327','A394','A40','A41','A427','B007','B377' |
| 2 | **Organ dysfunction** | 'D695','E872','G934','I46','I959','J80','J952','J96','K720','K729','N00',<br>'N17','R090','R092','R400','R401','R402','R41', 'R55', 'R57', 'R651', 'R572' |
| 3 | **Implicit Sepsis** | Organ dysfunction + Infection |
| 4 | **Infection** | 'A00','A01','A02','A03','A04','A05','A06','A07','A08','A09','A19',<br>'A20','A21','A22','A23','A24','A25','A26','A27','A28','A30','A31','A32','A36', 'A37','A38','A39','A42',<br>'A43', 'A44', 'A46','A48','A49','A54','A59','A690','A691','A699','A70','A74', 'A75',<br>'A77','A78','A79','A80','A81','A83','A84','A85','A86','A87',<br>'A88','A89','A90','A91','A92','A93','A94','A95','A96','A97','A98',<br>'A99','B00','B01','B02','B03','B04','B05','B06','B08','B09','B10','B25',<br>'B26','B27','B33','B34','B37','B38','B39','B40','B41','B42','B43','B44',<br>'B45','B46','B48','B49','B50','B54','B55','B57','B58','B59','B60','B64',<br>'B67','B95','B96','B97','B99','G00','G01','G02','G03','G04','G05','G06',<br>'G07','G08','H050', 'H602','H700','I00','I33','I38','I39','I400','J01','J02','J03',<br>'J04','J05','J06','J09','J10','J11','J12','J13','J14','J15','J16','J17','J18','J19','J20',<br>'J21','J22','J36','J390','J391','J85','J86','K35','K36','K37','K61','K630',<br>'K631','K65','K750', 'K810', 'K830','L02','L03','L030','L04','L08','M00','M01','M86','N10','N151',<br>'N30','N390','N410','N412','N413','N45','N70','N71','N72','N73','N74',<br>'N980','O030','O035','O045','O080','O23','O753',<br>'O85', 'O86','O883','O91','O98','R02','T802','T814','T826''T827','T835',<br>'T836','T845','T846','T847','T857','T880', 'U04', 'M726', 'N49', 'U071', 'U072' |
| 5 | **Cancer** | 'C00', 'C01', 'C02', 'C03', 'C04', 'C05', 'C06', 'C07', 'C08', 'C09', 'C10', 'C11', 'C12', 'C13', 'C14', 'C15',<br>'C16', 'C17', 'C18', 'C19','C20', 'C21', 'C22', 'C23', 'C24', 'C25', 'C26', 'C27', 'C28', 'C29',<br>'C30', 'C31', 'C32', 'C33', 'C34', 'C35', 'C36', 'C37', 'C38', 'C39', 'C40', 'C41', 'C42', 'C43', 'C44',<br>'C45', 'C46', 'C47', 'C48', 'C49','C50', 'C51', 'C52', 'C53', 'C54', 'C55', 'C56', 'C57', 'C58', 'C59',<br>'C60', 'C61', 'C62', 'C63', 'C64', 'C65', 'C66', 'C67', 'C68', 'C69', 'C70', 'C71', 'C72', 'C73', 'C74',<br>'C75', 'C76', 'C77', 'C78', 'C79','C80', 'C81', 'C82', 'C83', 'C84', 'C85', 'C86', 'C87', 'C88', 'C89',<br>'C90', 'C91', 'C92', 'C93', 'C94', 'C95', 'C96', 'C97', 'D32', 'D33', 'D35', 'D42', 'D43', 'D44', 'D45',<br>'D46', 'D47' |
| 6 | **Diabetes** | 'E10', 'E11', 'E12', 'E13', 'E14' |
| 7 | **Cardiovascular** | 'G45', 'H34', 'I00', 'I01', 'I02', 'I03', 'I04', 'I05', 'I06', 'I07', 'I08', 'I09', 'I10', 'I11', 'I12', 'I13', 'I14', 'I15',<br>'I16', 'I17', 'I18', 'I19', 'I20', 'I21', 'I22', 'I23', 'I24', 'I25', 'I26', 'I27', 'I28', 'I29', 'I30',<br>'I31', 'I32', 'I33', 'I34', 'I35', 'I36', 'I37', 'I38', 'I39', 'I40', 'I41', 'I42', 'I43', 'I44', 'I45', 'I46', 'I47',<br>'I48', 'I49', 'I50', 'I51', 'I52', 'I53', 'I54', 'I55', 'I56', 'I57', 'I58', 'I59', 'I60',<br>'I61', 'I62', 'I63', 'I64', 'I65', 'I66', 'I67', 'I68', 'I69', 'I70', 'I71', 'I72', 'I73', 'I74', 'I75', 'I76', 'I77',<br>'I78', 'I79', 'I80', 'I81', '182', 'I83', 'I84', 'I85', 'I86', 'I87', 'I88', 'I89', 'I90', 'I91', 'I92', 'I93',<br>'I94', 'I95', 'I96', 'I97', 'I98', 'I99' |
| 8 | **Lung** | 'J41', 'J42', 'J43', 'J44', 'J45', 'J46', 'J47', 'J84', 'J98' |
| 9 | **Dementia** | 'F00', 'F02', 'F03', 'G30', 'G31' |
| 10 | **Kidney** | 'N18' |
| 11 | **Liver** | 'K70', 'K72' |
| 12 | **Immune system** | 'D80', 'D81', 'D82', 'D83', 'D84', 'Z94' |

### Supplementary List 1. Groups of various microbiology tests

**'annet'**: ['ABSCESS', 'ABSCESS (TBA)', 'ABSCESS (VAB)', 'ACITES PÅ BL.K.FLASKE', 'AMPUTASJONSSTUMP', 'ANNET', 'ANNET (ANS)', 'ANNET (VAN)'],

**'anus'**: ['ANUSSEKRET', 'ANUSSEKRET (ANUM)', 'ANUSSEKRET (ANUP)', 'ANUSSEKRET (ANUS)', 'ANUSSEKRET (VANU)', 'ASCITES', 'ASPIRAT', 'ASPIRAT (VAS)', 'ASPIRAT PÅ BL.K.FLASKE', 'AUTOPSIMATRIALE', 'AUTOPSIMATRIALE (VAU)', 'AXILLE', 'AXILLE (MRSA)', 'BAKTERIESTAMME'],

**'bein'**: ['BEIN FRA BEINBANK', 'BEIN TIL BEINBANK', 'BEINMARG', 'BEINMARG (BEM)', 'BEINVEV'],

**'biopsi'**: ['BIHULESEKRET (BIH)', 'BIOPSI', 'BIOPSIMATERIALE', 'BIOPSIMATERIALE (TBI)', 'BIOPSIMATERIALE (VBI)'],

**'blod'**: ['BLOD - ISOLATOR', 'BLOD - ISOLATOR (TBLI)', 'BLODKULTUR (BLS)', 'BLODKULTUR (BLS1)'],

**'blood_culture_test': ['BLODKULTUR'],**

**'bronki'**: ['BRONKIALBØRSTE (VBR)', 'BRONKIALSKYLLEVÆSKE', 'BRONKIALSKYLLEVÆSKE (TBS)', 'BRONKIALSKYLLEVÆSKE (VBS)'],

**'melk'**: ['BRYSTMELK', 'BURSAVÆSKE', 'CERVIX (CERC)', 'CERVIX (CERM)', 'CERVIX-/URETHRASEKRET (CU', 'CERVIX-/VAGINALSEKRET', 'CERVIXSEKRET', 'CERVIXSEKRET (VCE)', 'CH-UROGENITALSEKRET', 'CH-UROGENITALSEKRET (UROM',

'CORNEAAVSKRAP', 'CYSTEINNHOLD', 'DIALYSAT PÅ BL.K.FLASKE', 'DIALYSEVÆSKE', 'DRENSPISS', 'DRENSVÆSKE', 'DRENSVÆSKE PÅ BL.K.FLASKE'],
**'edta'**: ['EDTA-BLOD', 'EDTA-BLOD (BEDT)', 'EDTA-BLOD (EDTA)', 'EDTA/UTSTRYK MALARIA', 'EJAKULAT (EJA)', 'EKSPEKTORAT', 'EKSPEKTORAT (EXS)', 'EKSPEKTORAT (TEX)', 'EKSPEKTORAT (VEK)', 'ELUAT', 'FISTEL', 'FOSTERVANN', 'FOSTERVANN (VFO)'],
**'faeces'**: ['FÆCES', 'FÆCES (FÆ)', 'FÆCES (FÆCB)', 'FÆCES (FÆCD)', 'FÆCES (FÆCP)', 'FÆCES (FÆD)', 'FÆCES (FÆFP)', 'FÆCES (FÆP)', 'FÆCES (VFÆ)', 'FÆCES (VFÆB)', 'FÆCES (VFÆN)', 'FÆCES (VFÆV)', 'GALLEVEISPRØVE', 'GENITALSEKRET (VGF)'],
**'hal'**: ['HALSSEKRET', 'HALSSEKRET (HALC)', 'HALSSEKRET (HALG)', 'HALSSEKRET (HALM)', 'HALSSEKRET (HAS)', 'HALSSEKRET (MRSA)', 'HALSSEKRET (VHA)', 'HALSSEKRET (VHAR)'],
**'hud'**: ['HUD', 'HUD (MRSA)', 'HUDAVSKRAP', 'HUDAVSKRAP (VHU)', 'HÅR', 'INDUSERT SPUTUM', 'INDUSERT SPUTUM (TSPU)', 'INNSTIKKSTED', 'KATETERSPISS', 'LARYNXSEKRET'],
**'led'**: ['LEDDVÆSKE', 'LEDDVÆSKE - ANRIKET', 'LGV (LYMFOGRAN. VENEREUM)', 'LYSAT', 'LYSKE (LYS)', 'MELK', 'MORSMELK'],
**'naso'**: ['MRSA REFERANSESTAMME (MRS', 'MUNNHULE', 'MUNNSEKRET', 'MUNNSEKRET (VMU)', 'NASOFARYNKS- OG HALSPRØVE', 'NASOPHARYNXASPIRAT (LUFA)', 'NASOPHARYNXASPIRAT (LUFT)', 'NASOPHARYNXASPIRAT (NAI)', 'NASOPHARYNXSEKRET', 'NASOPHARYNXSEKRET (NAS)', 'NASOPHARYNXSEKRET (VNAR)', 'NAVLESEKRET', 'NAVLESTRENG - BIT', 'NEGL', 'NEGL (NEGL)', 'NESESEKRET', 'NESESEKRET (DIA)', 'NESESEKRET (MRSA)'],
**'plasma'**: ['NONHUMANT MATERIALE', 'OPERASJONS-SÅR', 'OPPKAST', 'PACEMAKERTRÅD', 'PARAFININNSTØPT VEV', 'PD-DIALYSAT', 'PERICARDVÆSKE', 'PERICARDVÆSKE (VPEC)', 'PERINEUM', 'PERINEUM (MRSA)', 'PERITONEALVÆSKE', 'PERITONSILLÆRABSESS', 'PLACENTA', 'PLASMA', 'PLASMA (PLAS)', 'PLEURAV. PÅ BL.K.FLASKE', 'PLEURAVÆSKE', 'PLEURAVÆSKE (TPL)', 'PLEURAVÆSKE (VPLE)', 'PUSS (PSD)', 'PUSS/SEKRET', 'PUSS/SEKRET (TPU)', 'PUSS/SEKRET (VPUS)', 'RECTUMSEKRET (PREC)', 'SEKRET (SEK)', 'SERUM', 'SERUM (SSE)', 'SKYLLEVÆSKE (VSK)', 'SOPPKULTUR', 'SPINALVÆSKE', 'SPINALVÆSKE (VSP)', 'SPIRAL', 'SPISS AV CVK', 'SPUTUM (VSPU)', 'SÅRSEKRET', 'SÅRSEKRET (MRSA)', 'SÅRSEKRET (SÅS)', 'SÅRSEKRET (VSÅ)', 'TRACHEALASPIRAT', 'TRANSPLANTATMEDIUM', 'TRANSTRACHEALT ASPIRAT (T', 'TUBESEKRET (VTU)'],
**'tunge'**: ['TUNGESEKRET', 'TUNGESEKRET (TUNS)', 'TUNGESEKRET (VTUN)', 'TÅREVÆSKE'],
**'urin'**: ['URETHRA', 'URETHRA (UREM)', 'URETHRASEKRET', 'URETHRASEKRET (VUR)', 'URIN', 'URIN (CHLAMYDIA)', 'URIN (CHU)', 'URIN (CLUM)', 'URIN (CLUR)', 'URIN (TUR)', 'URIN (URS)', 'URIN (VURI)', 'URIN BLÆREPUNKSJON', 'URIN/TRANSPORTAGAR (URC)', 'UROGENITALSEKRET', 'UROGENITALSEKRET (URO)', 'USPESIFISERT (USP)', 'USPESIFISERT (VUS)', 'USPESIFISERT(MRSA)', 'UTERUSSEKRET (PUTE)', 'UTSTRYK', 'VAGINA', 'VAGINA (VAGM)', 'VAGINALPENSEL/URIN', 'VAGINALSEKRET', 'VAGINALSEKRET (CHP)', 'VAGINALSEKRET (VVG)', 'VESIKKELINNHOLD', 'VESIKKELINNHOLD (VESB)', 'VEV (TVV)', 'VEV/BIOPSI', 'VEV/BIOPSI (BIO)', 'VULVA (VVU)'],
**'ear'**: ['ØRESEKRET', 'ØRESEKRET (ØRS)', 'ØRESEKRET, HØYRE ØRE', 'ØRESEKRET, VENSTRE ØRE'],
**'eye'**: ['ØYEKAMMERVÆSKE', 'ØYESEKRET', 'ØYESEKRET (VØY)', 'ØYESEKRET (ØYC)', 'ØYESEKRET (ØYM)', 'ØYESEKRET - HØYRE ØYE', 'ØYESEKRET - HØYRE ØYE (VØ', 'ØYESEKRET - VENSTRE ØYE', 'ØYESEKRET - VENSTRE ØYE (' ]}

**Supplementary Table 9. Contaminant microbes:** List of microbes identified as contaminants

| | |
|---|---|
| **Contaminants** | 'BACILLUS CEREUS', 'STREPTOCOCCUS EQUI SSP EQUI', 'BACILLUS CIRCULANS', 'STREPTOCOCCUS EQUI SSP ZOOEPIDEMICUS', 'BACILLUS FIRMUS', 'STREPTOCOCCUS GORDONII', 'BACILLUS LICHENIFORMIS', 'STREPTOCOCCUS INTERMEDIUS', 'BACILLUS MEGATERIUM • STREPTOCOCCUS MITIS', 'BACILLUS PUMILUS', 'STREPTOCOCCUS MITIS', 'STREPTOCOCCUS ORALIS', 'BACILLUS SPECIES', 'STREPTOCOCCUS MUTANS', 'BACILLUS SPHAERICUS', 'STREPTOCOCCUS PYOGENES', 'BACILLUS SUBTILIS', 'STREPTOCOCCUS SALIVARIUS', 'COAGULASE NEGATIVE STAPHYLOCOCCUS', 'STREPTOCOCCUS SANGUINIS', 'CORYNEBACTERIUM JEIKEIUM', 'STREPTOCOCCUS VESTIBULARIS', 'CORYNEBACTERIUM SPECIES', 'STREPTOCOCCUS VIRIDANS GROUP', 'CORYNEBACTERIUM XEROSIS', 'MICROCOCCUS LUTEUS', 'MICROCOCCUS LYLAE', 'MICROCOCCUS LUTEUS', 'STAPHYLOCOCCUS ARLETTAE','MICROCOCCUS SPECIES', 'STAPHYLOCOCCUS CAPRAE', 'STAPHYLOCOCCUS SPP', 'STAPHYLOCOCCUS CARNOSUS SSP CARNOSUS', 'STAPHYLOCOCCUS AUREUS', 'STAPHYLOCOCCUS GALLINARUM', 'STAPHYLOCOCCUS AURICULARIS', 'STAPHYLOCOCCUS HOMINIS SSP HOMINIS','STAPHYLOCOCCUS CAPITIS', 'STREPTOCOCCUS ALACTOLYTICUS', 'STAPHYLOCOCCUS COHNII SSP COHNII', 'STREPTOCOCCUS CRISTATUS', 'STAPHYLOCOCCUS COHNII SSP UREALYTICUS', 'STREPTOCOCCUS CONSTELLATUS SSP CONSTELLATUS','STAPHYLOCOCCUS EPIDERMIDIS', 'STREPTOCOCCUS CONSTELLATUS SSP PHARYNGIS', 'STAPHYLOCOCCUS HAEMOLYTICUS', 'STREPTOCOCCUS HYOINTESTINALIS', 'STAPHYLOCOCCUS HOMINIS', 'STREPTOCOCCUS MITIS/STREPTOCOCCUS ORALIS', 'STAPHYLOCOCCUS INTERMEDIUS', 'STREPTOCOCCUS PARASANGUINIS', 'STAPHYLOCOCCUS KLOOSII', 'STREPTOCOCCUS PLURANIMALIUM', 'STAPHYLOCOCCUS LENTUS', 'STREPTOCOCCUS SOBRINUS', 'STAPHYLOCOCCUS LUGDUNENSIS','STREPTOCOCCUS THERMOPHILUS', 'STAPHYLOCOCCUS SACCHAROLYTICUS', 'STREPTOCOCCUS THORALTENSIS', 'STAPHYLOCOCCUS SAPROPHYTICUS', 'STREPTOCOCCUS SPP', 'STAPHYLOCOCCUS SCHLEIFERI', 'DIPHTHEROIDS SPP', 'STAPHYLOCOCCUS SCIURI', 'CORYNEBACTERIUM STRIATUM', 'STAPHYLOCOCCUS SIMULANS', 'NON HAEMOLYTIC STREPTOCOCCIS', 'STAPHYLOCOCCUS SPECIES', 'BABESIA SPP', 'STAPHYLOCOCCUS WARNERI', 'CORYNEBACTERIUM MINUTISSMUM', 'STAPHYLOCOCCUS XYLOSUS','CORYNEBACTERIUM AMYCOLATUM', 'STREPTOCOCCUS AGALACTIAE', 'MICROMONAS MICROS', 'STREPTOCOCCUS ANGINOSUS', 'STAPHYLOCOCCUS PASTEURI', 'STREPTOCOCCUS CONSTELLATUS' |
